# Assessment of the Effects of a Sage (*Salvia officinalis*) Extract on Cognitive Performance in Adolescents and Young Adults

**DOI:** 10.1101/2021.05.28.21257776

**Authors:** Kieron D. Edwards, Anne Dubberke, Nadin Meyer, Simone Kugel, Juliane Hellhammer

## Abstract

**Background:** Cognitive health is a major concern for many people, and with potential benefits to academic and professional life, maximising cognitive performance is of interest far beyond the older demographics. Several natural products have been suggested as nootropics, including the herb sage. Previous assessments of various *Salvia* species have reported a range of effects on cognitive performance and mood in both older adult and younger adult populations. This study was conducted with Sibelius™: Sage, an aqueous-ethanol extract of *S. officinalis*, to assess for the beneficial effects on cognitive performance in adolescents (12-14 year olds) and young adults (18-25 year olds).

**Methods and Findings:** An acute, double**-**blind, placebo-controlled study was conducted with two single doses of Sibelius™: Sage (150 mg and 300 mg). Cognitive performance was evaluated using CogTrack™, which probes aspects of cognitive performance covering attention, working memory and episodic/declarative memory through a series of computer-based tasks. Consistent with previous study of Sibelius™: Sage a significant effect was seen on the Immediate Word Recall task in young adults; suggesting acute treatment benefits to verbal episodic/declarative memory performance. Physiological effects of the treatment on salivary cortisol and oxytocin levels, as well as blood pressure and heart rate were also assessed, with limited evidence of an effect on these factors. No adverse events or side-effects linked to the study product intake was observed. The study was registered at the German Clinical Trials Register (DRKS-ID: DRKS00015716).

**Conclusions:** A significant improvement due to the Sage extract was shown for a task assessing short-term episodic memory (Immediate Word Recall), supporting beneficial effects on cognitive performance in young adults that are consistent with previous reports in healthy older adults. These findings suggest that further investigation of the effects observed in this study in larger, long-term human volunteer studies could be beneficial to pursue.

## Introduction

Cognition broadly covers the mental actions of acquiring, processing, understanding, storing, and recalling information or knowledge. Whilst some facets of cognitive ability are maintained or even improve with age – such as general knowledge and wisdom – other facets – such as attention, working memory, and episodic memory – show declines during what is considered normal ageing [1]. Coupled with global trends of ageing populations [2], and the increased incidence and prevalence of cognitive diseases, such as dementia and Alzheimer’s Disease (AD) [3], this has resulted in growing awareness of the need to support and maintain cognitive health and performance as we age. However, cognitive performance is not only an issue for older individuals or those suffering with cognitive diseases. Changes to working practices, such as longer working hours, are placing increasing pressures on professional performance [4], so cognitive benefits are also being actively sought by healthy individuals, not just diseased [5], and in younger demographics.

Given the diversity and breadth of secondary metabolites present in the plant kingdom, some of which have the potential to interact with and modulate elements of human biology including the central nervous system [6], natural products provide opportunities for cognitive-enhancing supplements (which are also often referred to as nootropics). Based on long-term use in traditional medicine or epidemiological data, the nootropic potential of multiple ingredients such as *Ginkgo biloba* [7], *Bacopa monnieri* [8], and *Curcuma longa* [9] have been considered.

The herb sage is another botanical that shows strong potential for beneficial effects on cognitive performance [10]. The *Salvia* genus is part of the *Lamiaceae* family, which contains a large number of species, although two have been most extensively studied for cognitive effects: *S. officinalis* (Common Sage) and *S. lavandulifolia* (Spanish Sage). Along with other *Salvia*, both of these species are proposed to improve cognitive performance through modulation of the metabolism of the important memory neurotransmitter acetylcholine. These cholinergic effects are specifically through inhibition of the enzymes Acetylcholinesterase (AChE) and Butylcholinesterase (BChE), which have been demonstrated multiple times in *Salvia* extracts [11–14]. Monoterpenes, including 1,8-cineole, have been proposed as strong candidates for mediating these activities [12], although phenolic acids such as rosmarinic acid present in sage also display the same activities [15]. AChE inhibiting activity is also shared by the front-line treatments of symptoms in mild-to-moderate AD. This includes Donepezil, Rivastigmine and Galanthamine, which have been shown to slow the decline in cognition, function, and behaviour of AD patients [16]. Indeed, based on this activity, sage has been proposed as a potential treatment for AD [17], with studies of an aqueous-ethanol extract of *S. officinalis* showing promising results [18].

In addition to cholinergic activity, the anti-oxidant and anti-inflammatory effects of sage [19], alongside neurotropic effects, have been proposed to contribute towards beneficial cognitive and anxiolytic effects of the plant (reviewed in [10]). Human volunteer studies have previously shown significant effects on cognitive performance in healthy [13] and cognitively diseased [18] older adults, as well as younger adults [11,12,14,20]. Within healthy individuals, these studies suggested that acute cognitive benefits are often seen within one hour of a single dose, and maintained for several hours; with the major benefits in the areas of episodic memory, attention, and to a lesser extent working memory [11–14,20].

Although of the same genus, direct equivalence of *S. officinalis* and *S. lavandulifolia* cannot necessarily be assumed given biochemical differences between the plants [21]. In addition to this, studies in *S. lavandulifolia* have typically been conducted using essential oils [12,14,20], whereas studies in *S. officinalis* have focussed on whole leaf [11] or aqueous-ethanol extracts [13], which will have very different biochemical profiles. It has been proposed that *S. lavandulifolia* essential oil may be preferable to *S. officinalis* essential oil due to low-to-non-existent levels of the neurotoxic compounds α-and β-thujone [12]. However, it is possible to generate *S. officinalis* extracts that are low in thujone, and a comparison of the studies using similar assessment approaches support comparatively strong benefits to cognitive performance achieved with *S. officinalis* [13].

This current study was undertaken to test the acute effect on cognitive performance in adolescents and younger adults of a standardised, low-thujone extract of *S. officinalis* (Sibelius™: Sage); developed utilising Sibelius’ Chronoscreen™ platform. This extract was previously shown to have significant acute benefits in a double-blind, randomised, placebo-controlled, cross-over study in healthy older adults [13], and we now show evidence for cognitive effects in younger adults consistent with other studies of sage extracts [11,12,14,20].

## Methods

### Participants

The study was conducted in two separate cohorts: adolescents (aged 12-14 years), and young adults (aged 18-25 years). All participants (and their legal representatives in the case of the adolescent cohort) first received information on study background and procedure during a telephone call. Participants meeting all inclusion criteria and none of the exclusion criteria (see Supplementary methods file S5) were invited for a first visit at the study site. Extensive study information was handed out and a written informed consent was obtained from all participants (or their legal representatives in the case of the adolescent cohort) prior to initiation of the study. 37 adolescents were recruited to the study with 36 completing it, and 36 young adults were recruited with all completing the study. A summary of the study participants, including descriptive statistics for demography and other baseline characteristics, is available in Supplementary methods file S5.

The study was performed in accordance with the ethical principles originating in the Declaration of Helsinki and consistent with the Good Clinical Practice guidelines of the International Conference on Harmonization and applicable regulatory requirements on bioethics. The study protocol was submitted on August 08, 2018 and approved on September 26, 2018 by the Ethics Commission of the State Chamber of Medicine of Rhineland-Palatinate, Deutschhausplatz 3, 55116 Mainz, Germany under the processing number 2018-13597. The study was registered at the German Clinical Trials Register (DRKS-ID: DRKS00015716).

### Cognitive measures

Cognitive performance was evaluated using CogTrack™ [22], which is based on the sensitive tests in the Cognitive Drug Research System (CDR) that has been the most widely used system of its type in worldwide clinical research and was used in previous assessment of the sage extract in older adults [13]. Tests of CogTrack™ were used in the study to assess i) attention, concentration and vigilance (simple reaction time, digit vigilance, and choice reaction time tasks), ii) working memory and executive control (spatial working memory, and numeric working memory tasks), and iii) episodic/declarative memory (verbal recall and recognition, and picture recognition tasks). A brief description of each of the tasks is provided in Supplementary methods file S5. Scores for the individual task outcomes were also collapsed into the cognitive factors secondary memory, working memory, speed of memory, accuracy of attention, and speed of attention as described previously [13] and included in exploratory analysis for this study.

### Physiological assessments

#### Salivary cortisol

Salivary cortisol levels [23] were measured to assess the Cortisol Awakening Response (CAR) and the reactivity of the hypothalamic-pituitary-adrenal (HPA)-axis in response to the stimulus of the cognitive testing. Nine saliva samples were collected in total, with the first three being collected by participants at home immediately upon awakening and then at 30 minutes and 45 minutes post-awakening to assess the CAR. These samples were taken using Salivettes^®^ (Sarstedt, Nuembrecht, Germany) by placing a synthetic swab in the mouth, moving it gently for approx. one minute and then replacing the swab back in the plastic tube. The remaining six samples were collected at the study site before and after each cognitive assessment on the test day, with samples collected by spitting into a plastic tube without using a swab to obtain whole saliva. Saliva samples collected at home were cooled and delivered at site, all saliva samples were stored at −20°C until analysis.

Salivary cortisol levels were determined in duplicates at daacro’s Saliva Lab Trier (Germany) using a high sensitivity salivary cortisol competitive immunoassay kit [24,25] manufactured by Salimetrics (Carlsbad, USA). The kit contains a 96-well microplate coated with monoclonal antibodies to cortisol. Briefly, on the day of assay, the saliva samples were thawed, vortexed and centrifuged at 1500 x g for 15 minutes to remove mucins and other particulate matter. After assay completion, optical density was read on a BioTek ELx808 microplate reader at 450 nm (with correction at 490 nm). The calculations for determining the cortisol concentration on each sample were carried out following the kit manufacturer’s instructions and using the Gen 5 v.2.04 software which enabled, as required, a 4-parameter non-linear regression standard curve to be generated. Assay quality was measured by calculating the intra-assay coefficient of variation. Inter-assay variability was checked with the low and high controls on each microplate. The intra-and inter-assay coefficients of variation were 3.40 % and 4.86 %, respectively.

#### Salivary oxytocin

For the assessment of oxytocin, whole saliva was collected in a plastic tube at the study site as per the method described for salivary cortisol at 35 minutes prior to the study product administration and 80 minutes after the study product administration. Saliva samples were stored at site at −20°C and then analyzed for oxytocin levels by radioimmunoassay at RIAgnosis (Sinzing, Germany). In brief, for each sample 300 μl of saliva was evaporated (SpeedVac, Thermoscentific Inc, Waltham, USA) and 50 μl of assay buffer was added followed by 50 μl antibody (raised in rabbits against oxytocin). After a 60-min pre-incubation interval, 10 µL 125I-labeled tracer (PerkinElmer, Waltham, USA) was added, and samples were allowed to incubate for 3 days at 4°C. Unbound radioactivity was precipitated by activated charcoal/Dextran (Sigma–Aldrich, St Louis, USA). Under these conditions, an average of 50 % of total counts are bound with <5 % non-specific binding. The detection limit is in the 0.1-0.5 pg/sample range. The intra-and inter-assay variabilities were <10 %. Generally, all samples from an individual challenge are assayed in the same batch. Serial dilutions of saliva samples containing high levels of endogenous oxytocin run strictly parallel to the standard curve indicating immuno-identity.

#### Vital parameters and body composition related measurements

Blood pressure (BP; systolic and diastolic) and heart rate (HR) were assessed by using an automated blood pressure measurement device (OMRON M10-IT, OMRON Medizintechnik Handelsgesellschaft mbH, Gottlieb-Daimler-Str. 10, 68165 Mannheim). If blood pressure measures were below the normal range (systolic < 100, diastolic < 60), a second measurement was performed and recorded in the paper Case Report Form which, if also below the normal range, would lead to study exclusion. Body Mass Index (BMI), Body Fat Percentage (BFP) and Muscle Percentage (MP) were assessed by using a body composition monitor (BF 511 Körperanalyse-Monitor, OMRON Medizintechnik Handelsgesellschaft mbH, Gottlieb-Daimler-Str.10, 68165 Mannheim). Measurements of BMI, BFP, MP, BP and pulse were recorded at visit 1 (V1), and during the test day (V3) BP and heart rate were obtained after arrival and before and after each test session.

### Other variables

#### Psychometric assessments

Prior to the test day, during visit 2 (V2), psychometric assessment of the participants was undertaken. The young adult cohort undertook the Trier Inventory for Chronic Stress (TICS) [26]. Stress chronicity is measured by the frequency of stressful events perceived retrospectively within the last three months. Answers are given on a five-point rating scale, where 0 resembles “never” and 4 “very often”. For analysis, the questionnaire items are assigned to 10 scales: Work overload, Social overload, Pressure to succeed, Work dissatisfaction, Excessive demands at work, Lack of social recognition, Social stress, Social isolation and Chronic worrying; the last scale presents a Screening Scale for Chronic Stress. The adolescent cohort undertook the SSKJ 3-8 R questionnaire [27], which is applicable from the third to the eighth grade and covers the vulnerability to potential stressors, the existing potential for coping with stress, and the potential physical and psychological symptoms associated with stress.

#### Sleep quality, lifestyle parameters, and development

Participants were asked to fill out a questionnaire related to their lifestyle and sleep habits. On a scale from 0 to 10, they indicated how much fruits, vegetables, meat, fish, and sweets and unhealthy snacks they consumed (0 = none at all, 10 = multiple portions daily). On a scale from 0 to 10, they further rated their level of activity (0 = no physical exercise, 10 = several hours of physical exercise daily). With regard to sleep, participants indicated whether and how often they woke up during the preceding night of V2 and V3, how long they slept and their perception of recovery after awaking, again on a scale from 0 to 10 (0 = not recovered, 10 = very much recovered).

Prior to the test day, during visit 2, adolescent participants were also assessed via the Pubertal Development Scale, which is a self-report instrument that measures levels of pubertal maturation in adolescence into five different pubertal stages [28]

### Treatments

The active treatment for the study was a standardised aqueous-ethanolic extract of *Salvia officinalis* (sage) provided by Sibelius Ltd. Sage plant material of known and invariant provenance was grown in the United Kingdom, according to defined production protocols and Good Agricultural Practice standards. Sage leaves were dried in an artificially heated (gas fired) hot air drier at temperatures less than 70°C and then soaked in ethanol (68 % w/w) for 48h. The resultant solution was then concentrated using a climbing film evaporator and dried in a vacuum oven to a final ratio of approximately 7.5:1 dry plant material to extract. The resulting material was milled using a 45 mesh to produce the final Sibelius™:Sage extract, which is a commercial extract standardised to a minimum level of 2.5 % w/w rosmarinic acid (batch number 44878 used in this study).

Test products were packed and labelled according to the randomization plan by the pharmacy of the University of Mainz (Apotheke der Universitätsmedizin der Johannes Gutenberg-Universität Mainz). 150 mg of placebo (99.5 % mannitol, 0.5 % aerosil), and 150 mg or 300 mg of *S. officinalis* extract were packed into size 0, opaque gelatine capsules for use in the study. The label contained information on product name, study code, storage conditions, manufacturer of the test product, sponsor of the clinical trial, study site, best before date and ingredients of the test product.

### Procedure

This study was a single-center, double-blind, controlled, parallel group design study with three arms and a total duration of three visits per participant (V1, V2, and V3). Subjects for the adolescent study were randomly assigned to one of two experimental groups (150 mg *Salvia officinalis* L. or control product) and subjects for the young adult study were randomly assigned to one of the three experimental groups (300 mg *Salvia officinalis* L., 150 mg *Salvia officinalis* L. or control product). Participants were randomised into treatment groups sequentially during enrolment based on a blind randomisation sequence generate with the package blockrand (Snow, 2013) in R (Version 3.5.1).

Details of the three visits are available in the Supplementary methods file S5 and also described briefly below. Visit 1 (V1) followed telephone screening as was conducted to assess inclusion/exclusion criteria and inform eligible participants of requirements for the study. During Visit 2 (V2) participants were asked to re-confirm their medical history, intake of concomitant medication, their sleep duration and quality, their lifestyle and level of activity as well as their alcohol consumption. Participants also filled out the TICS assessments and underwent two training sessions for familiarization with the CogTrack™ cognitive tests. Participants were instructed not to consume any alcohol, and that they should not consume anything but water for one hour before V3, which was scheduled on the next day.

On the morning of Visit 3 (V3) participants were asked to collect saliva samples with Salivettes^®^ as described above to assess the CAR. V3 took place between 12 noon and 3pm (starting time), approximately 24 hours after V2. Participants were given a detailed introduction to the study procedure of V3, and asked about their medical history, intake of concomitant medication, their sleep duration and quality, and their alcohol consumption. Vital signs (BP, HR) were assessed. Participants were instructed to drink only water and not to eat during V3 besides the standard meal provided. At 35 minutes prior to product intake (−35 minutes), participants BP and HR were assessed, and saliva sample #1 was collected. Participants also completed SAM #1 and VAS #1. Baseline cognitive tests were then performed on CogTrack™ at −25 minutes and following this, saliva sample #2 was collected, participants BP and HR were assessed, and participants completed SAM #2 and VAS #2 at −5 minutes.

Product administration occurred according to the randomisation plan, and then cognitive tests were performed (CogTrack™) at 60 minutes post study product administration and 150 minutes post study product administration. Saliva samples were collected, BP and HR were assessed just prior to and just after each cognitive test session (+50 minutes, +80 minutes, + 140 minutes, and +170 minutes). Subjects also received a standardised meal at +90 minutes.

### Statistical analysis

Data for the two cohorts were analysed separately. Accuracy in the CogTrack™ Picture Recognition task for the adolescent cohort was analyzed for the intention to treat (ITT) and per protocol populations. All other outcomes were analyzed for the ITT population, while safety outcomes were analyzed for the safety population. No correction for multiple testing was performed. Two-sided hypothesis testing (α = .05) was performed. All calculations were performed with version 3.5 of the statistic program R.

All data is descriptively summarized in tabular form by treatment group and assessment time in Supplementary data files S1 and S2. Statistical outliers were assessed and excluded from analyses. Statistical outliers were defined as values of three standard deviations below or above the mean for univariate values or a Mahalanobis distance significant on the *P* < 0.001 level according to the corresponding degrees of freedom as a multivariate outlier for normally distributed data. For non-normally distributed data any data point more than 1.5 interquartile ranges below the first quartile or above the third quartile was defined as univariate outlier.

Results from the CogTrack™ cognitive testing were analyzed using linear mixed models including time (three measuring points, categorical) and group (two treatment groups) as fixed effects and a random intercept for participants. Covariates were tested in a model only including time and entered in the final model, if the *p*-value was below 0.1. If 2 covariates exhibited co-linearity with each other, alternative models including only one were build and the best model according to Akaike information criterion chosen.

Main effects for time and group (i.e. significant change over time, and overall group differences) as well as time × group interaction effects (i.e. different changes over time with respect to group) were assessed using F-tests. These F-tests only reveal general significant effects for any change in time, any group differences, and any time × group interaction. In case of a main effect for time, Tukey pairwise comparisons were performed to reveal which time points significantly differed from each other. For a significant interaction effect and a significant main effect of group, contrast analyses were performed to identify significant differences in the change of the outcome over time points between treatment groups (comparing the active treatment group to the placebo group).

For the biomarker endpoints (HR, BP and salivary cortisol), if there were no systematic differences in these endpoints before and after a CogTrack™ cognitive testing, then they were analyzed using linear mixed models with time as a continuous predictor, potential covariates, the factor group (two treatment groups) and time × group interaction effects. If there were systematic differences in these endpoints before and after CogTrack™ cognitive testing, two (pre-resp. post-cognitive testing) linear mixed models including time and group (two treatment groups) as fixed effects and a random intercept for participants were build.

## Results and Discussion

### Treatment effect on cognitive measures

This study was undertaken to assess whether intake of an *S. officinalis* extract leads to an increase of cognitive performance including episodic memory, working memory, attention, concentration and inhibition processes compared to the placebo group in adolescents or young adults. Results from the assessment of cognitive performance by CogTrack™, as well as for physiological outcomes, are summarised in Supplementary data Tables S1 and S2 for the adolescent and young adult cohorts respectively, and a summary of the statistical analyses can be found in Supplementary data S3 and S4 for the adolescent and young adult cohorts respectively.

Previous assessment of the same *S. officinalis* extract in a placebo-controlled study in older adults showed strong positive effects on various aspects of cognitive performance; with particular benefits on episodic/declarative memory [13]. As such, it was anticipated that the current study would show beneficial effects on tests related to this aspect of cognitive performance. Episodic memory was assessed by tests with either visual (Picture Recognition Accuracy) or verbal (Word Recall Accuracy) stimuli. A significant effect of the *S. officinalis* treatment was shown for performance of the Immediate Word Recall Accuracy test in the young adult cohort for both the 150 mg and 300 mg doses of the extract at the 1 hour post-treatment test time-point (Supplementary data file S4).

Figure 1 summarises the change in Immediate Word Recall Accuracy performance from baseline performance across the two post-treatment test time-points for the young adults. The data shows that a decrease in accuracy at the second CogTrack™ testing, 1 hour post-product intake, shown in the placebo group was attenuated by the intake of either the 150 mg or 300 mg dose of *S. officinalis* extract (Figure 1). A significant effect was seen on this same test in a previous study of older adults treated with the same *S. officinalis* extract; which also showed the same pattern in fall-off in performance between baseline testing and later tests for the placebo [13] (as have other studies using equivalent methodology [20]).

**Figure 1.**
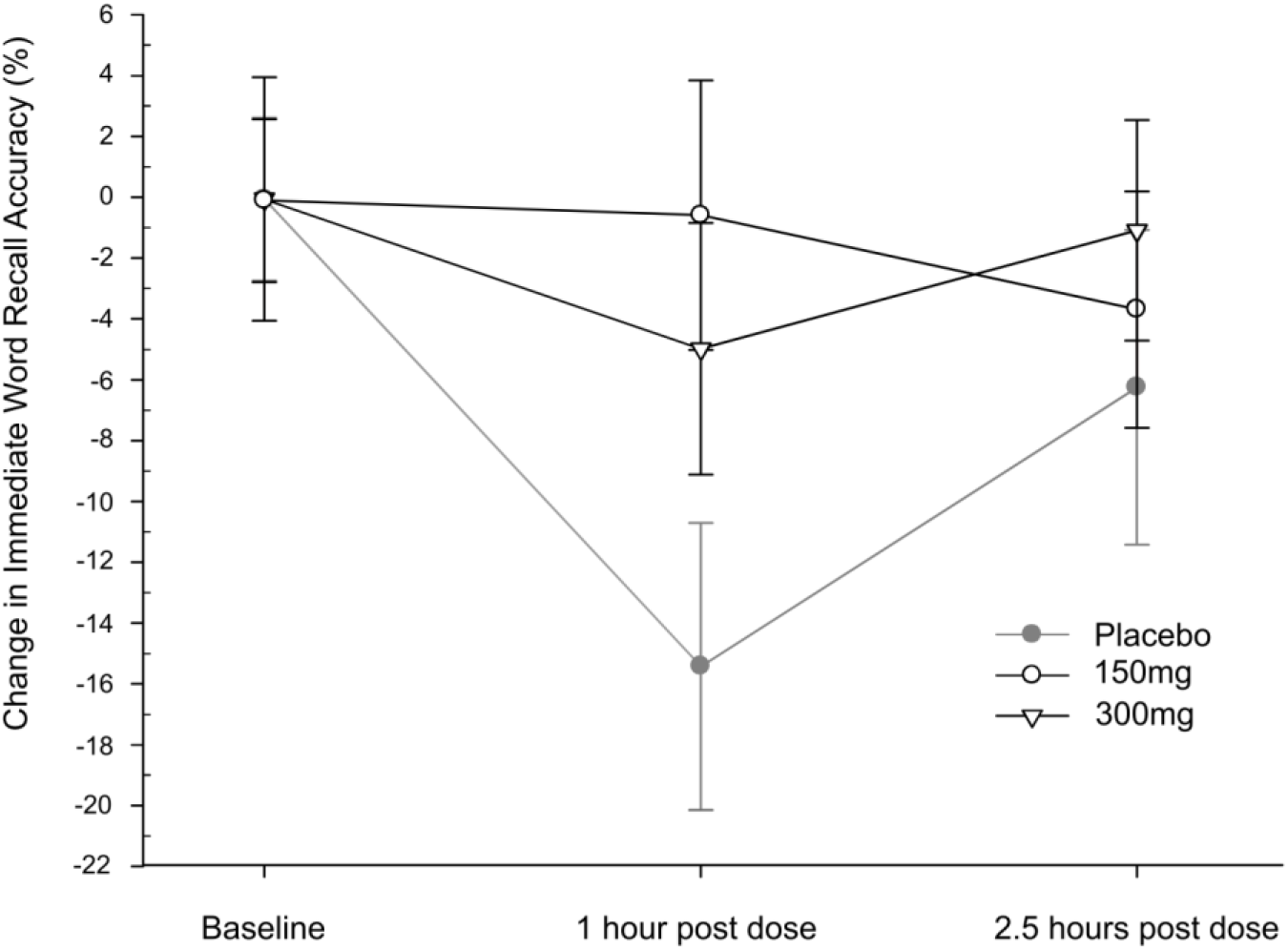
Effect of *S. officinalis* extract on Immediate Word Recall Accuracy in young adults. Line plot showing the change in Immediate Word Recall Accuracy % in the young adult population at the 1 hour and 2.5 hours post-treatment time points compared to performance during baseline testing. Data for the placebo (closed grey circles), 150 mg *S. officinalis* extract (open black circles) and 300 mg *S. officinalis* extract (open black triangles) treatment arms are represented, following removal of outliers as described in the method section. Error bars show Standard Error of the Mean.

Immediate Word Recall Accuracy performance appears to decline with age; with baseline scores in the range of *ca*. 50-60 % in this study (Supplementary Table S2 A), versus *ca*. 30-35 % previously shown in older adults [13]. A similar age-related decline is also observed for Picture recognition accuracy, but in this case performance levels beyond 50 years of age are similar to those observed in this study (*ca*. 80-90% accuracy; Supplementary Table S2 A) and don’t really start to show strong declines until ages of above 60 years [22]. High baseline performance in this task might suggest that the potential to see beneficial effects in younger cohorts as assessed in this study was limited, whereas there was still capacity to see improvements to the word recall accuracy task given the relatively lower general performance level. Furthermore, whereas the Sage treatment attenuated the fall-off in Immediate Word Accuracy performance between base-line and the 1 hour post-treatment time-point, as observed in the placebo arm for the young adults (a response which as noted above has now been observed across multiple studies [13,20]), no such fall-off in performance was observed for the picture recognition task (Supplementary Table S2). Similarly, the adolescent group, who unlike the young adults did not show a significant effect for the Immediate Word Recall task, also did not show this same fall-off in performance between the baseline and later time-points for this task (Supplementary Table S1). This suggests that cognitive benefits may manifest themselves differently between different task types and age-groups.

In addition to demonstrating beneficial effects of the *S. officinalis* extract on the same test in an independent study of older adults [13], improved performance of both doses of the *S. officinalis* extract on Immediate Word Recall Accuracy in this study is in line with benefits of the herb on cognitive function in young and healthy adults for a different set of cognitive tests in a previous study based on ingestion of dried *S. officinalis* leaf [11]. It is also broadly consistent with positive cognitive effects on cognition in young adults for extracts from *S. lavandulifolia*, including benefits to the same Immediate Word Recall Accuracy task as observed in this study [12,14,20].

Previous assessment of the Sage extract in older adults showed broader benefits to cognitive outcomes [13]. This may be due to well-known cognitive performance [1] and physiological changes [29] associated with age, meaning that adolescents and young adults are much less likely than older adults to show deficits that can easily be compensated by a single administration of a nutritional supplement. However, acute cognitive benefits have been demonstrated in young adults in this study, as well as in multiple other studies from single doses of *Salvia* extracts or leaf [11,12,14,20].

The results of this study and previous assessment of the *S. officinalis* extract [13] do appear to suggest a possible difference in the timing of the cognitive benefits between younger and older adults, with a more acute benefit in younger adults on the Immediate Word Recall Accuracy test (present only at the 1 hour post-treatment time-point; Supplementary Table S2 A and Figure 1) and a later onset but more prolonged effect in older adults (present at the 2.5 hours and 4 hours post-treatment time-points [13]). This timing difference may be due to different mechanisms or age-related physiological differences, but it also might indicate different rates of metabolism of active constituents within the extract. Consistent with this, studies of acute cognitive effects for *S. lavandulifolia* essential oil in young adults [12,14,20] tended to show early and less prolonged effects; although it was suggested that this depends on the type of cognitive task with early benefits (*ca*. 1 hour post-treatment) shown for attention or simple memory tasks, and later benefits (*ca*. 4 hours post-treatment) shown for more complex mental tasks and fatigue [12]. Assuming similar mechanisms are involved in the cognitive benefits for both *Salvia* extract types, then this suggests the possibility that the 2.5 hours post-treatment time-point utilised in this study was not optimal to see maximal cognitive benefits: Too late to capture the early benefits and too early to capture the later benefits.

Taking this further, it is interesting to consider that visual episodic memory tests may require application of a broader set of neurocognitive skills, such as encoding not only the visual stimulus of the object in question but also spatial information, as compared to verbal episodic memory tests [30]. Given the dynamics of cognitive benefits for *Salvia* extracts suggested above, this might explain why a significant effect is seen on the Immediate Word Recall Accuracy test at 1 hour, but not on the putatively more complex Picture Recognition Accuracy test, which may instead have shown benefits but at later time-points to those assessed in this study.

Another possible explanation is that the dose used for the adolescents was not in the beneficial therapeutic window of the *S. officinalis* extract. The importance of an adequate dose is stressed by dose-dependent effects detected by Tildesley *et al*. for *S. lavandulifolia*, where both their lowest and highest dose did not result in improved cognitive function, while effects were found for two doses in between [14]. As no reference work existed for the present age group of 12-14 year olds, further experimentation with different doses might be of value.

It is well known that during the age of puberty, the brain undergoes substantial changes [31]. It can therefore be expected that participants vary largely with regard to their performance on cognitive tasks already at baseline. Furthermore, a number of confounding factors were recorded for both age cohorts in this study and used in the statistical models to reduce overall variance. In particular, demographics such as weight and body-fat-percentage as well as indicators of chronic stress (Trier Inventory for Chronic Stress [TICS] and Cortisol Awakening Response [CAR]), diet-related factors, and quality and length of sleep in the night before the testing proved to be influential with regard to the cognitive performance (see individual tables in Supplementary data S3 and S4).The previous studies showing broader cognitive benefits of *Salvia* extracts were based on cross-over designs, whereas the current study was conducted as a parallel design, which may therefore have required a larger number of subjects to be tested than anticipated during the design of this study given the resultant baseline variation observed between subjects and number of significant confounding factors.

Some support is given to the importance of confounding factors such as sleep, which is well known to influence cognitive performance [32]. Exploratory analyses with triple interaction terms showed two significant interactions with sleep quality in the adolescent cohort: Higher sleep quality in the 150 mg *S. officinalis* treatment group was associated with a higher increase in perceived exhaustion over time as well as a higher increase in accuracy of secondary memory from baseline to 1 hour post-treatment compared to placebo; a feature of memory that was also significantly improved in older adults previously [13]. It must be noted that these analyses are very exploratory, and given the small sample size they are based, on could easily be the result of chance. However, they do point to potentially interesting interactions that could be explored further in future studies.

Whether the test product can be expected to have a similar effect in all participants despite the different initial situation is an open question, and points in the possible direction of individual needs and a personalized nutritional profile followed by supplementation, respectively. Here, especially stress concepts might help further exploring beneficial effects of sage by studying a young study population characterized by a pronounced psychobiological burden of stress.

### Treatment effects on physiological measures

To consider potential physiological effects of the *S. officinalis* extract on participants, assessment of heart rate, blood pressure and salivary levels of oxytocin and cortisol were taken during the test day (Supplementary data S1 B and S2 B). The *S. officinalis* extract showed no significant effects on diastolic or systolic blood pressure in either the adolescent or young adult cohorts. Similarly no treatment effect was shown for heart rate in the adolescent cohort. A statistically significant effect was observed for heart rate in the young adult group (Supplementary data file S4), driven by the 150 mg treatment group showing a lower decrease in heart rate versus the placebo (Supplementary data file S4, Supplementary Table S2 B and Figure 2). However, the 150 mg treatment group measured lower heart rate at baseline when compared to the placebo or 300 mg treatment groups. Therefore, the observed effect is likely due to a regression to the mean, as heart rate for the 150mg treatment group subsequently approached mean measurements of the other two treatments during the course of the testing session (Figure 2). Taken together, the data would suggest that the *S. officinalis* does not alter blood pressure or heart rate, which differentiates it from other well-known nootropic compounds such as caffeine [33] that have been linked to stimulatory effects on these factors [34].

**Figure 2.**
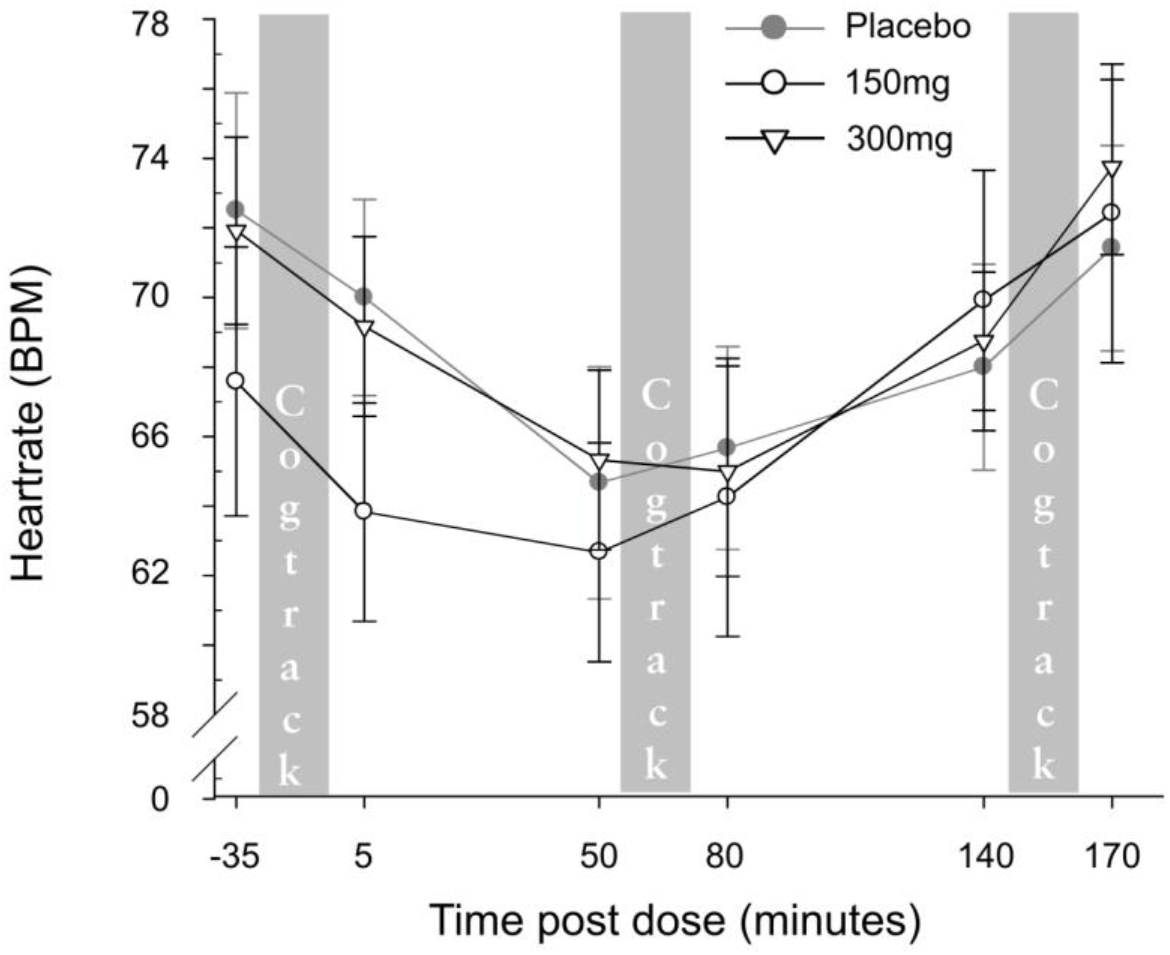
Effect of *S. officinalis* extract on heart rate in young adults. Line plots showing the effect of *S. officinalis* extract on heart rate in the young adult study population. Data for the placebo (closed grey circles), 150 mg *S. officinalis* extract (open black circles) and 300 mg *S. officinalis* extract (open black triangles) treatment arms are represented, following removal of outliers as described in methods. Assessments were taken before and after baseline, 1 hour post-treatment and 2.5 hours post-treatment CogTrack™ cognitive assessment (marked with grey bars in the plots). Error bars show Standard Error of the Mean.

Caffeine has also been linked to increased levels of cortisol [35]. Corticoids play an important regulatory role in the carbohydrate, lipid and protein metabolism and therefore in energy supply to the brain, as well as having immunosuppressive and anti-inflammatory properties. Activation of a hormonal cascade by the hypothalamic-pituitary-adrenal (HPA)-axis by stressors ultimately results in the release of cortisol from the cortex of the suprarenal gland, and measurements of HPA-axis activity and reactivity are frequently used as markers in stress research [36,37].Whereas HPA-activity varies considerably during the day [38], the Cortisol Awakening Response (CAR) is a physiological reaction to awakening [39], and is characterized by an increase in cortisol around 30-45 minutes after awakening with peak values rising by 50-156% [40,41]. The magnitude of the CAR reflects several psychological and physiological manifestations including chronic stress and burnout [42].

As noted above, the magnitude of participants CAR, assessed in the morning ahead of the test day, as well as other subjective psychometric measurements related to stress and anxiety, had a significant effect on several cognitive outcomes during the statistical analysis (Supplementary data S3 and S4). However, there was no significant effect of *S. officinalis* extract on HPA-axis activity as measured by salivary cortisol levels during the test day in either cohort. Similarly, there was no significant treatment effect in either cohort on the salivary levels of oxytocin, a brain neuropeptide that is a regulator of anxiety, stress-coping, and sociality [43], which effectively acts a counterpart to cortisol. Finally, there were no adverse in the adolescent cohort and only one mild adverse event recorded in the young adult cohort within the 150 mg treatment group and deemed unlikely to be related to the product intake, which supports the safety of the ingredient up to 150 mg and 300 mg for the 12-14 years and 18-25 years cohorts respectively.

## Conclusions

Sibelius ™: Sage, an aqueous-ethanolic extract of *S. officinalis* had previously shown acute benefits to cognitive performance in older adults [13]. The results of this study support cognitive benefits for this same extract in younger adults. In particular both tested doses of the extract showed an attenuation of fall-off in performance in an Immediate Word Recall Accuracy task 1 hour post intake of the supplement by young adults (aged 18-25). Beneficial effects of the extract on this same task have now been shown in two independent populations, as well as in other studies assessing the acute cognitive benefits of *Salvia* extracts [14,20], which offers support of this being a real effect.

Multiple confounding factors were discovered with significant effects on the study outcomes during the analysis. Knowledge of these factors can be used to inform the design of future studies to better account for their effects; in particular potential consideration of cross-over designs, as used in the other studies of *Salvia* extracts [11–14,20], or increased sample size. Also, given the potential of beneficial cognitive effects manifesting in significant improvements to different types cognitive tasks with different temporal dynamics, consideration of tasks and time-points to optimally assess these outcomes should be given. Certainly, consideration for a longer-term study should be given, as a single dose of a dietary supplement may not be optimal to show strong significant benefits, particularly in healthy younger populations with probably less scope to display beneficial effects. Furthermore, a longer study would also allow investigation of other mechanisms, such as anti-inflammatory effects, that could contribute towards long-term cognitive benefits.

Finally, in addition to results from previous assessment [13], the lack of adverse events recorded in this study offers support for the safety of the extract of at least up to 150 mg for adolescents and 300 mg for young adults.

## Data Availability

Data are not available on-line. Reasonable requests will be considered.

## Acknowledgements

Special thanks to Dominik Seithel for data management, Christof Zintel for programing the online diary and Helma Veerman for her relentless telephone screening. Finally, we wish to extend our gratitude to all the study participants who made this study possible.

## Competing and Conflicting Interests

This study was fully sponsored by Sibelius Limited, 20 East Central, 127 Olympic Avenue, Milton Park, Abingdon, Oxfordshire, OX14 4SA, United Kingdom. The sponsors were involved in discussion of the design of the study as well as in the writing of the paper.

### Abbreviations

AD: Alzheimer’s Disease
AChE: Acetylcholinesterase
BChE: Butylcholinesterase
BP: Blood Pressure
BFP: Body Fat Percentage
BMI: Body Mass Index
CAR: Cortisol Awakening Response
HR: Heart Rate
(HPA)-axis: hypothalamic-pituitary-adrenal
ITT: Intention To Treat
MP: Muscle Percentage
PP: Per Protocol
SAM: Self Assessment Manakin
TICS: Trier Inventory for Chronic Stress
VAS: Visual Analogue Scale

## Author Contributions

Kieron Edwards: Conceptualization, Methodology, Writing and editing of manuscript, Visualization, Project Administration.

Anne Dubberke: Conceptualization, Methodology, Writing and editing of manuscript. Nadin Meyer: Study Management, Writing and editing of manuscript.

Simone Kugel: Study Management, Writing and editing of manuscript.

Juliane Hellhammer: Conceptualization, Methodology, Resources, Writing and editing of manuscript, Supervision.

## List of Supplementary Files

File S1 – Summary results tables for outcomes in adolescents.

File S2 - Summary results tables for outcomes in young adults.

File S3 – Summary analysis tables for outcomes in adolescents

File S4 - Summary analysis tables for outcomes in young adults

File S5 – Supplementary methods

