## Supplementary Table S1 for "Assessment of the Effects of a Sage (*Salvia officinalis*) Extract on Cognitive Performance in Adolescents and Young Adults"

| | | Baseline | | 1h post treatment | | | 2.5h post treatment | | |
|---|---|---|---|---|---|---|---|---|---|
| | | Mean | SD | Mean | Delta to baseline | SD | Mean | Delta to baseline | SD |
| Picture Recognition accuracy (%) | Placebo | 84.17 | 13.58 | 79.33 | -4.84 | 9.14 | 82.83 | -1.34 | 11.83 |
| | 150mg | 80.16 | 8.92 | 75.47 | -4.69 | 8.43 | 77.5 | -2.66 | 9.17 |
| Picture Recognition Speed (msec) | Placebo | 1155.03 | 163.46 | 1128.2 | -26.83 | 165.72 | 1111.26 | -43.77 | 149.47 |
| | 150mg | 1174.19 | 209.11 | 1222.1 | 47.91 | 250.56 | 1170.13 | -4.06 | 280.34 |
| Word Recognition Accuracy (%) | Placebo | 80 | 14.28 | 79.44 | -0.56 | 11.1 | 76.85 | -3.15 | 12.5 |
| | 150mg | 78.73 | 9.73 | 75.18 | -3.55 | 13.05 | 74.44 | -4.29 | 9.43 |
| Word Recognition Reaction Time (msec) | Placebo | 882.59 | 130.33 | 932.24 | 49.65 | 319.83 | 920.47 | 37.88 | 352.77 |
| | 150mg | 855.69 | 162.06 | 876.47 | 20.78 | 211.69 | 898.57 | 42.88 | 218.53 |
| Immediate Word Recall Accuracy (%) | Placebo | 43.33 | 12.37 | 39.26 | -4.07 | 10.7 | 40 | -3.33 | 13.91 |
| | 150mg | 41.25 | 10.05 | 40.74 | -0.51 | 8.52 | 36.3 | -4.95 | 14.68 |
| Immediate Word Recall Errors | Placebo | 0.33 | 0.59 | 0.17 | -0.16 | 0.38 | 0.39 | 0.06 | 0.7 |
| | 150mg | 0.53 | 0.7 | 0.22 | -0.31 | 0.55 | 0.56 | 0.03 | 0.86 |
| Delayed Word Recall Accuracy (%) | Placebo | 30.74 | 14.67 | 21.85 | -8.89 | 20 | 26.67 | -4.07 | 11.88 |
| | 150mg | 31.73 | 12.86 | 22.96 | -8.77 | 14.63 | 21.85 | -9.88 | 14.78 |
| Delayed Word Recall Errors | Placebo | 0.67 | 0.59 | 0.61 | -0.06 | 0.92 | 0.56 | -0.11 | 0.98 |
| | 150mg | 0.95 | 1.39 | 1.06 | 0.11 | 0.94 | 1.06 | 0.11 | 1.26 |
| Numeric Working Memory Accuracy (%) | Placebo | 89.44 | 7.69 | 91.3 | 1.86 | 8.49 | 91.11 | 1.67 | 7.14 |
| | 150mg | 93.33 | 5.67 | 94.07 | 0.74 | 4.05 | 94.63 | 1.3 | 5.38 |
| Numeric Working Memory Reaction Time (msec) | Placebo | 911.51 | 236.72 | 832.91 | -78.6 | 171.29 | 815.4 | -96.11 | 182.16 |
| | 150mg | 811.16 | 133.79 | 819.41 | 8.25 | 152.26 | 801.02 | -10.14 | 155.48 |
| Spatial Working Memory Accuracy (%) | Placebo | 94.62 | 4.99 | 94.62 | 0 | 6.52 | 93.68 | -0.94 | 7 |
| | 150mg | 96.32 | 3.19 | 95.49 | -0.83 | 3.94 | 94.76 | -1.56 | 5.29 |
| Spatial Working Memory Reaction Time (msec) | Placebo | 777.39 | 209.51 | 760.18 | -17.21 | 196.71 | 712.75 | -64.64 | 157.67 |
| | 150mg | 729.71 | 128.79 | 716.29 | -13.42 | 197.12 | 687.21 | -42.5 | 130.42 |
| Simple Reaction Time (msec) | Placebo | 358.6 | 34.2 | 357.4 | -1.2 | 36.55 | 365.66 | 7.06 | 36.97 |
| | 150mg | 350.49 | 35.88 | 357.2 | 6.71 | 32.21 | 357.02 | 6.53 | 42.71 |
| Digit Vigilance Targets Detected (%) | Placebo | 94.32 | 4.39 | 93.46 | -0.86 | 5.98 | 93.46 | -0.86 | 6.62 |
| | 150mg | 95.32 | 4.74 | 94.45 | -0.87 | 4.71 | 93.21 | -2.11 | 5.93 |
| Digit Vigilance Reaction Time (msec) | Placebo | 508.6 | 37.06 | 505.65 | -2.95 | 25.16 | 516.2 | 7.6 | 47.95 |
| | 150mg | 501.06 | 36.82 | 503.73 | 2.67 | 37.14 | 496.35 | -4.71 | 39.58 |
| Digit Vigilance False Alarms | Placebo | 1.56 | 1.79 | 2.11 | 0.55 | 2.63 | 1.44 | -0.12 | 1.62 |
| | 150mg | 1.42 | 1.3 | 2 | 0.58 | 1.64 | 3.11 | 1.69 | 4.64 |
| Choice Reaction Time Accuracy (%) | Placebo | 93.44 | 3.68 | 95.11 | 1.67 | 2.76 | 95.33 | 1.89 | 3.07 |
| | 150mg | 93.26 | 3.78 | 93.89 | 0.63 | 4.92 | 92.56 | -0.7 | 5.77 |
| Choice Reaction Time (msec) | Placebo | 465.87 | 38.02 | 478.36 | 12.49 | 47.53 | 485.7 | 19.83 | 55.83 |
| | 150mg | 483.24 | 54.65 | 480.04 | -3.2 | 61.65 | 482.24 | -1 | 74.41 |
| Secondary Memory Accuracy (range: 0 – 400) | Placebo | 239.77 | 38.44 | 221.67 | -18.1 | 27.66 | 226.3 | -13.47 | 32.34 |
| | 150mg | 233.82 | 29.72 | 216.39 | -17.43 | 29.62 | 211.87 | -21.95 | 30.97 |
| Working Memory Accuracy (range: 0 – 200) | Placebo | 184.06 | 10.03 | 185.91 | 1.85 | 13.91 | 184.79 | 0.73 | 12.61 |

| | | | | | | | | | |
|---|---|---|---|---|---|---|---|---|---|
| | 150mg | 189.65 | 7.52 | 189.56 | -0.09 | 5.47 | 189.39 | -0.26 | 6.06 |
| Speed of Memory (msec) | Placebo | 3726.52 | 570.62 | 3653.52 | -73 | 554.58 | 3559.88 | -166.64 | 645.29 |
| | 150mg | 3570.75 | 486.61 | 3634.27 | 63.52 | 678.01 | 3556.93 | -13.82 | 649.25 |
| Accuracy of Attention (%) | Placebo | 93.88 | 3.11 | 94.28 | 0.4 | 3.28 | 94.4 | 0.52 | 3.98 |
| | 150mg | 94.29 | 3.51 | 94.17 | -0.12 | 3.47 | 92.88 | -1.41 | 4.05 |
| Speed of Attention (msec) | Placebo | 1333.07 | 86.72 | 1341.41 | 8.34 | 94.6 | 1367.56 | 34.49 | 124.81 |
| | 150mg | 1334.78 | 107.57 | 1340.96 | 6.18 | 105.85 | 1335.62 | 0.84 | 128.15 |
| Picture Recognition Original Stimuli Accuracy (%) | Placebo | 85.83 | 12.98 | 82.5 | -3.33 | 10.61 | 85.28 | -0.55 | 11.44 |
| | 150mg | 83.68 | 10.39 | 80 | -3.68 | 11.25 | 81.94 | -1.74 | 12.5 |
| Picture Recognition New Stimuli Accuracy (%) | Placebo | 85.56 | 14.84 | 79.72 | -5.84 | 10.07 | 80.28 | -5.28 | 11.94 |
| | 150mg | 80.53 | 11.29 | 75 | -5.53 | 10.71 | 76.61 | -3.92 | 9.15 |

**Table S1 A – Table of cognitive outcome measures in adolescents**

Table summarising the adolescent subjects' performance on the computer battery of tests. Test outcomes are presented on rows, separated by treatment group. Mean scores and standard deviation (SD) are presented at the baseline, 1 hour post treatment, and 2 hour post treatment assessment time-points. The difference in score to the baseline is also presented for the 1 hour post-treatment, and 2 hour post-treatment assessment time-points.

| | | Pre-baseline | | Post-baseline | | Pre-1h | | Post-1h | | Pre-2.5h | | Post-2.5h | |
|---|---|---|---|---|---|---|---|---|---|---|---|---|---|
| | | Mean | SD | Mean | SD | Mean | SD | Mean | SD | Mean | SD | Mean | SD |
| Heart rate (bpm) | Placebo | 70.67 | 9.79 | 69.78 | 13.74 | 64.89 | 9.78 | 66.22 | 9.6 | 71.06 | 11.09 | 73.33 | 13.89 |
| | 150mg | 73.32 | 13.7 | 70.68 | 11.85 | 66.11 | 12.32 | 66.56 | 11.92 | 70.78 | 13.47 | 71.83 | 12.4 |
| Diastolic blood pressure (mmHG) | Placebo | 67.17 | 13.22 | 63.72 | 8.68 | 63.5 | 7.5 | 66 | 7.47 | 67.33 | 11.09 | 64.89 | 8.63 |
| | 150mg | 65.89 | 6.74 | 62.42 | 8.76 | 67.33 | 5.52 | 66.78 | 6.25 | 65.72 | 7.44 | 66.89 | 8.59 |
| Systolic blood pressure (mmHG) | Placebo | 108.44 | 13.66 | 106.28 | 10.31 | 107.28 | 7.23 | 108.83 | 8.3 | 110.39 | 12 | 112 | 12.31 |
| | 150mg | 106.79 | 8.42 | 105.68 | 9.69 | 105.67 | 9.59 | 104.5 | 8.4 | 108.22 | 12.44 | 108.22 | 7.64 |
| Cortisol (nmol/l) | Placebo | 2.82 | 0.99 | 2.38 | 0.75 | 2.26 | 0.92 | 2.58 | 1.19 | 3.74 | 2.05 | 3.35 | 2.22 |
| | 150mg | 2.92 | 1.13 | 2.71 | 1 | 2.22 | 1.13 | 2.54 | 1.23 | 3.28 | 2.4 | 2.86 | 1.44 |
| Oxytocin (pg/ml) | Placebo | 1.6 | 0.45 | | | | | 1.43 | 0.43 | | | | |
| | 150mg | 1.66 | 0.39 | | | | | 1.52 | 0.37 | | | | |

**Table S1 B – Table of physiological outcomes in adolescents**

Table summarising the physiological assessments for the adolescent subjects. Test outcomes are presented on rows, separated by treatment group. Mean scores and standard deviation (SD) are presented for assessment time-points pre- and post- the baseline, 1 hour post-treatment, and 2 hour post-treatment assessment time-points for each of the measurements aside from oxytocin, which was only assessed at baseline and 1-hour post treatment time-points.
