## Supplementary Data S3 for "Assessment of the Effects of a Sage (*Salvia officinalis*) Extract on Cognitive Performance in Adolescents and Young Adults"

### 3 Supplementary Data File S3

#### 3.1 Baseline characteristics and control variables

**Table 1: Inferential statistics for baseline characteristics and control variables**

| **F-tests for continuous control variables** | | | | |
|---|---|---|---|---|
| Control variable | **Numerator DF** | **Denominator DF** | **F-value** | **p-value** |
| PDS | 1 | 35 | 0.304 | 0.585 |
| SSKJ 3-8 R psychological stress symptoms | 1 | 35 | 0.058 | 0.811 |
| subscale anger | 1 | 35 | 1.014 | 0.321 |
| subscale sadness | 1 | 35 | 1.057 | 0.311 |
| subscale anxiety | 1 | 35 | 0.560 | 0.459 |
| subscale wellbeing | 1 | 35 | 1.146 | 0.292 |
| CAR increase | 1 | 35 | 3.111 | 0.087 |
| Sleep quality | 1 | 35 | 0.390 | 0.536 |
| Sleep duration | 1 | 35 | 0.300 | 0.587 |
| Fruit and vegetable intake | 1 | 35 | 0.468 | 0.498 |
| Sweets intake | 1 | 35 | 0.100 | 0.754 |
| Level of activity | 1 | 35 | 0.020 | 0.890 |
| Weight | 1 | 35 | 2.328 | 0.136 |
| BMI | 1 | 35 | 1.720 | 0.198 |
| WHR | 1 | 35 | 0.191 | 0.664 |
| BFP | 1 | 35 | 0.421 | 0.521 |
| MP | 1 | 35 | 0.088 | 0.768 |
| **Fisher's exact test for categorical control variables** | | | | |
| | | | | **p-value** |
| Gender | | | | 0.915 |

#### 3.2 Cognitive outcomes

**Table 2: Inferential statistics for picture recognition accuracy in the ITT population, outcome was square transformed**

| | lme model summary | | | |
|---|---|---|---|---|
| | **Estimate** | **t-value** | **DF** | **p-value** |
| Intercept (placebo, baseline) | 6022.543 | 8.09 | 67 | <.001 |
| 1hr post dose | -1037.1 | -2.65 | 67 | 0.010 |
| 2.5hr post dose | -735.713 | -1.88 | 67 | 0.065 |
| 150 mg group | -782.388 | -1.57 | 33 | 0.125 |
| Gender | 650.633 | 1.55 | 33 | 0.131 |
| Sweets intake | 207.634 | 2.08 | 33 | 0.046 |
| 1hr post dose:150 mg | 334.774 | 0.61 | 67 | 0.543 |
| 2.5hr post dose:150 mg | 310.582 | 0.57 | 67 | 0.572 |
| | **F-tests** | | | |
| | **Numerator DF** | **Denominator DF** | **F-value** | **p-value** |
| Intercept (placebo, baseline) | 1 | 67 | 1252.24 | <.001 |
| Time | 2 | 67 | 5.13 | 0.008 |
| Group | 1 | 33 | 2.99 | 0.093 |
| Gender | 1 | 33 | 3.18 | 0.084 |
| Sweets intake | 1 | 33 | 4.30 | 0.046 |
| Time:group | 2 | 67 | 0.232 | 0.794 |
| | **Post-hoc Tukey tests for time** | | | |
| | **Estimate** | **z-value** | | **p-value** |
| 1hr post dose - baseline | -863.3 | -3.195 | | 0.004 |
| 2.5hr post dose – baseline | -574.0 | -2.125 | | 0.085 |
| 2.5hr post dose - 1hr post dose | 289.3 | 1.076 | | 0.529 |

**Table 3: Inferential statistics for picture recognition accuracy in the PP population, outcome was square transformed**

| | lme model summary | | | |
|---|---|---|---|---|
| | **Estimate** | **t-value** | **DF** | **p-value** |
| Intercept (placebo, baseline) | 7066.03 | 8.08 | 58 | <.001 |
| 1hr post dose | -884.583 | -2.01 | 58 | 0.049 |
| 2.5hr post dose | -264.167 | -0.6 | 58 | 0.550 |
| 150 mg group | -603.544 | -1.11 | 27 | 0.275 |
| Gender | 867.752 | 1.93 | 27 | 0.065 |
| SSKJ 3-8 R | -105.085 | -1.84 | 27 | 0.076 |
| Palliative emotion regulation | | | | |
| 1hr post dose:150 mg | 147.083 | 0.24 | 58 | 0.811 |
| 2.5hr post dose:150 mg | -150.286 | -0.25 | 58 | 0.807 |
| | **F-tests** | | | |
| | **Numerator DF** | **Denominator DF** | **F-value** | **p-value** |
| Intercept (placebo, baseline) | 1 | 58 | 1015.80 | <.001 |
| Time | 2 | 58 | 3.53 | 0.036 |
| Group | 1 | 27 | 3.47 | 0.074 |
| Gender | 1 | 27 | 3.49 | 0.073 |
| SSKJ 3-8 R | 1 | 27 | 3.39 | 0.076 |
| Palliative emotion regulation | | | | |
| Time:group | 2 | 58 | 0.12 | 0.889 |
| | **Post-hoc Tukey tests for time** | | | |
| | **Estimate** | **z-value** | | **p-value** |
| 1hr post dose - baseline | -808.7 | -2.685 | | 0.020 |
| 2.5hr post dose – baseline | -341.7 | -1.135 | | 0.493 |
| 2.5hr post dose - 1hr post dose | 466.9 | 1.550 | | 0.268 |

**Table 4: Inferential statistics for picture recognition reaction time in the ITT population, outcome was log transformed**

| | **lme model summary** | | | |
|---|---|---|---|---|
| | **Estimate** | **t-value** | **DF** | **p-value** |
| Intercept (placebo, baseline) | 7.042 | 173.45 | 67 | <.001 |
| 1hr post dose | -0.024 | -0.8 | 67 | 0.425 |
| 2.5hr post dose | -0.038 | -1.28 | 67 | 0.206 |
| 150 mg group | 0.012 | 0.2 | 35 | 0.840 |
| 1hr post dose:150 mg | 0.055 | 1.33 | 67 | 0.188 |
| 2.5hr post dose:150 mg | 0.005 | 0.11 | 67 | 0.914 |
| | **F-tests** | | | |
| | **Numerator DF** | **Denominator DF** | **F-value** | **p-value** |
| Intercept (placebo, baseline) | 1 | 67 | 74404.60 | <.001 |
| Time | 2 | 67 | 2.08 | 0.133 |
| Group | 1 | 35 | 0.36 | 0.550 |
| Time:group | 2 | 67 | 1.08 | 0.345 |

**Table 5: Inferential statistics for word recognition accuracy in the ITT population**

| | lme model summary | | | |
|---|---|---|---|---|
| | **Estimate** | **t-value** | **DF** | **p-value** |
| Intercept (placebo, baseline) | 32.61 | 2.18 | 67 | 0.033 |
| 1hr post dose | -0.555 | -0.25 | 67 | 0.803 |
| 2.5hr post dose | -3.147 | -1.42 | 67 | 0.161 |
| 150 mg group | -1.743 | -0.51 | 33 | 0.611 |
| PDS | 4.936 | 2.93 | 33 | 0.006 |
| SSKJ 3-8 R Psychological stress symptoms wellbeing | 4.344 | 2.44 | 33 | 0.02 |
| 1hr post dose:150 mg | -1.604 | -0.51 | 67 | 0.613 |
| 2.5hr post dose:150 mg | -1.063 | -0.34 | 67 | 0.735 |

| | F-tests | | | |
|---|---|---|---|---|
| | **Numerator DF** | **Denominator DF** | **F-value** | **p-value** |
| Intercept (placebo, baseline) | 1 | 67 | 2976.85 | <.001 |
| Time | 2 | 67 | 2.85 | 0.065 |
| Group | 1 | 33 | 0.56 | 0.458 |
| PDS | 1 | 33 | 7.2 | 0.011 |
| SSKJ 3-8 R Psychological stress symptoms wellbeing | 1 | 33 | 5.95 | 0.020 |
| Time:group | 2 | 67 | 0.13 | 0.875 |

**Table 6: Inferential statistics for word recognition reaction time in the ITT population**

| | lme model summary | | | |
|---|---|---|---|---|
| | **Estimate** | **t-value** | **DF** | **p-value** |
| Intercept (placebo, baseline) | 828.229 | 9.01 | 57 | <.001 |
| 1hr post dose | -38.359 | -1.71 | 57 | 0.092 |
| 2.5hr post dose | -49.461 | -2.26 | 57 | 0.028 |
| 150 mg group | 4.966 | 0.14 | 29 | 0.892 |
| BFP | -5.959 | -2.88 | 29 | 0.007 |
| Fruit and vegetable intake | -24.925 | -3 | 29 | 0.006 |
| Sweets intake | 15.315 | 1.84 | 29 | 0.075 |
| Sleep duration | 25.538 | 3.28 | 29 | 0.003 |
| 1hr post dose:150 mg | 32.644 | 1.04 | 57 | 0.304 |
| 2.5hr post dose:150 mg | 75.777 | 2.44 | 57 | 0.018 |
| | **F-tests** | | | |
| | **Numerator DF** | **Denominator DF** | **F-value** | **p-value** |
| Intercept (placebo, baseline) | 1 | 57 | 3007.75 | <.001 |
| Time | 2 | 57 | 1.05 | 0.358 |
| Group | 1 | 29 | <.01 | 0.979 |
| BFP | 1 | 29 | 6.65 | 0.015 |
| Fruit and vegetable intake | 1 | 29 | 7.14 | 0.012 |
| Sweets intake | 1 | 29 | 4.60 | 0.041 |
| Sleep duration | 1 | 29 | 10.87 | 0.003 |
| Time:group | 2 | 57 | 2.98 | 0.059 |

**Table 7: Inferential statistics for immediate word recall accuracy in the ITT population**

| | lme model summary | | | |
|---|---|---|---|---|
| | **Estimate** | **t-value** | **DF** | **p-value** |
| Intercept (placebo, baseline) | 32.983 | 6.81 | 68 | <.001 |
| 1hr post dose | -4.074 | -1.29 | 68 | 0.202 |
| 2.5hr post dose | -3.333 | -1.05 | 68 | 0.295 |
| 150 mg group | -1.536 | -0.41 | 34 | 0.685 |
| Gender | 7.762 | 2.57 | 34 | 0.015 |
| 1hr post dose:150 mg | 3.347 | 0.75 | 68 | 0.455 |
| 2.5hr post dose:150 mg | -1.836 | -0.41 | 68 | 0.681 |
| | **F-tests** | | | |
| | **Numerator DF** | **Denominator DF** | **F-value** | **p-value** |
| Intercept (placebo, baseline) | 1 | 68 | 840.63 | <.001 |
| Time | 2 | 68 | 1.78 | 0.176 |
| Group | 1 | 34 | 0.31 | 0.583 |
| Gender | 1 | 34 | 6.60 | 0.015 |
| Time:group | 2 | 68 | 0.69 | 0.504 |

**Table 8: Inferential statistics for immediate word recall errors in the ITT population**

| | glm model summary | | |
|---|---|---|---|
| | **Estimate** | **z-value** | **p-value** |
| Intercept (placebo, baseline) | -0.23033 | -0.274 | 0.784 |
| 1hr post dose | -0.77678 | -0.863 | 0.388 |
| 2.5hr post dose | 0.00007 | 0.000 | 1.000 |
| 150 mg group | 0.48650 | 0.565 | 0.572 |
| CAR increase | -0.09924 | -1.505 | 0.132 |
| 1hr post dose:150 mg | -0.75833 | -0.610 | 0.542 |
| 2.5hr post dose:150 mg | -0.14956 | -0.134 | 0.893 |
| | **LR-tests** | | |
| | **DF** | **LR Chisquare-value** | **p-value** |
| Time | 2 | 3.8546 | 0.146 |
| Group | 1 | 0.1609 | 0.688 |
| CAR increase | 1 | 2.2655 | 0.132 |
| Time:group | 2 | 0.3945 | 0.821 |

**Table 9: Inferential statistics for delayed word recall accuracy in the ITT population**

| | lme model summary | | | |
|---|---|---|---|---|
| | Estimate | t-value | DF | p-value |
| Intercept (placebo, baseline) | 4.764 | 0.55 | 68 | 0.585 |
| 1hr post dose | -8.89 | -2.81 | 68 | 0.006 |
| 2.5hr post dose | -4.075 | -1.29 | 68 | 0.202 |
| 150 mg group | 2.273 | 0.56 | 33 | 0.577 |
| Sleep quality | 1.563 | 1.52 | 33 | 0.139 |
| PDS | 4.897 | 2.6 | 33 | 0.014 |
| 1hr post dose:150 mg | -0.103 | -0.02 | 68 | 0.982 |
| 2.5hr post dose:150 mg | -6.029 | -1.35 | 68 | 0.18 |

| | F-tests | | | |
|---|---|---|---|---|
| | Numerator DF | Denominator DF | F-value | p-value |
| Intercept (placebo, baseline) | 1 | 68 | 278.05 | <.001 |
| Time | 2 | 68 | 9.05 | 0.003 |
| Group | 1 | 33 | 0.11 | 0.740 |
| Sleep quality | 1 | 33 | 4.87 | 0.034 |
| PDS | 1 | 33 | 6.75 | 0.014 |
| Time:group | 2 | 68 | 1.19 | 0.309 |

| | Post-hoc Tukey tests for time | | |
|---|---|---|---|
| | Estimate | z-value | p-value |
| 1hr post dose - baseline | -9.010 | -4.034 | < 0.001 |
| 2.5hr post dose – baseline | -7.157 | -3.205 | 0.004 |
| 2.5hr post dose - 1hr post dose | 1.852 | 0.826 | 0.687 |

**Table 10: Inferential statistics for delayed word recall errors in the ITT population, outcome was log transformed after adding 1 to each value**

| | lme model summary | | | |
|---|---|---|---|---|
| | **Estimate** | **t-value** | **DF** | **p-value** |
| Intercept (placebo, baseline) | 0.642 | 4.71 | 60 | <.001 |
| 1hr post dose | -0.093 | -0.84 | 60 | 0.406 |
| 2.5hr post dose | -0.231 | -2.00 | 60 | 0.050 |
| 150 mg group | -0.176 | -1.22 | 34 | 0.231 |
| CAR increase | -0.02 | -2.06 | 34 | 0.047 |
| 1hr post dose:150 mg | 0.378 | 2.34 | 60 | 0.023 |
| 2.5hr post dose:150 mg | 0.319 | 1.90 | 60 | 0.062 |
| | **F-tests** | | | |
| | **Numerator DF** | **Denominator DF** | **F-value** | **p-value** |
| Intercept (placebo, baseline) | 1 | 60 | 57.91 | <.001 |
| Time | 2 | 60 | 2.24 | 0.115 |
| Group | 1 | 34 | 1.25 | 0.271 |
| CAR increase | 1 | 34 | 4.59 | 0.040 |
| Time:group | 2 | 60 | 3.11 | 0.052 |

**Table 11: Inferential statistics for numeric working memory accuracy in the ITT population**

| | lme model summary | | | |
|---|---|---|---|---|
| | **Estimate** | **t-value** | **DF** | **p-value** |
| Intercept (placebo, baseline) | 87.811 | 34.64 | 65 | <.001 |
| 1hr post dose | 4.277 | 2.49 | 65 | 0.016 |
| 2.5hr post dose | 2.93 | 1.73 | 65 | 0.088 |
| 150 mg group | 4.117 | 2.43 | 32 | 0.021 |
| BFP | -0.199 | -2.89 | 32 | 0.007 |
| Fruit and vegetable intake | 0.56 | 2.05 | 32 | 0.048 |
| Sweets intake | 0.694 | 2.54 | 32 | 0.016 |
| 1hr post dose:150 mg | -3.463 | -1.45 | 65 | 0.150 |
| 2.5hr post dose:150 mg | -1.56 | -0.66 | 65 | 0.511 |
| | **F-tests** | | | |
| | **Numerator DF** | **Denominator DF** | **F-value** | **p-value** |
| Intercept (placebo, baseline) | 1 | 65 | 31889.29 | <.001 |
| Time | 2 | 65 | 2.49 | 0.091 |
| Group | 1 | 32 | 4.61 | 0.039 |
| BFP | 1 | 32 | 9.84 | 0.004 |
| Fruit and vegetable intake | 1 | 32 | 3.44 | 0.073 |
| Sweets intake | 1 | 32 | 6.33 | 0.017 |
| Time:group | 2 | 65 | 1.06 | 0.353 |

Table 12: Inferential statistics for numeric working memory reaction time in the ITT population

| | lme model summary | | | |
|---|---|---|---|---|
| | Estimate | t-value | DF | p-value |
| Intercept (placebo, baseline) | 748.454 | 6.20 | 68 | <.001 |
| 1hr post dose | -78.599 | -2.40 | 68 | 0.019 |
| 2.5hr post dose | -96.103 | -2.93 | 68 | 0.005 |
| 150 mg group | -63.332 | -1.17 | 32 | 0.251 |
| Fruit and vegetable intake | -19.796 | -1.63 | 32 | 0.113 |
| Sweets intake | 20.397 | 1.70 | 32 | 0.099 |
| SSKJ 3-8 R Stress vulnerability | 15.713 | 2.04 | 32 | 0.050 |
| 1hr post dose:150 mg | 89.498 | 1.94 | 68 | 0.057 |
| 2.5hr post dose:150 mg | 88.612 | 1.92 | 68 | 0.059 |
| | **F-tests** | | | |
| | **Numerator DF** | **Denominator DF** | **F-value** | **p-value** |
| Intercept (placebo, baseline) | 1 | 68 | 1326.58 | <.001 |
| Time | 2 | 68 | 2.58 | 0.083 |
| Group | 1 | 32 | 0.87 | 0.357 |
| Fruit and vegetable intake | 1 | 32 | 3.66 | 0.065 |
| Sweets intake | 1 | 32 | 3.61 | 0.067 |
| SSKJ 3-8 R Stress vulnerability | 1 | 32 | 4.11 | 0.051 |
| Time:group | 2 | 68 | 2.48 | 0.091 |

**Table 13: Inferential statistics for spatial working memory accuracy in the ITT population**

| | lme model summary | | | |
|---|---|---|---|---|
| | **Estimate** | **t-value** | **DF** | **p-value** |
| Intercept (placebo, baseline) | 107.517 | 12.32 | 64 | 0 |
| 1hr post dose | 1.36 | 1.21 | 64 | 0.231 |
| 2.5hr post dose | 1.006 | 0.88 | 64 | 0.383 |
| 150 mg group | 2.531 | 1.86 | 31 | 0.072 |
| WHR | -21.517 | -2.04 | 31 | 0.05 |
| CAR increase | 0.187 | 2.07 | 31 | 0.047 |
| SSKJ 3-8 R stress coping strategies: seeking social support | 0.211 | 1.64 | 31 | 0.111 |
| 1hr post dose:150 mg | -2.386 | -1.53 | 64 | 0.132 |
| 2.5hr post dose:150 mg | -1.961 | -1.23 | 64 | 0.222 |
| | **F-tests** | | | |
| | **Numerator DF** | **Denominator DF** | **F-value** | **p-value** |
| Intercept (placebo, baseline) | 1 | 64 | 37846.11 | <.0001 |
| Time | 2 | 64 | 0.02 | 0.9763 |
| Group | 1 | 31 | 0.44 | 0.5112 |
| WHR | 1 | 31 | 3.73 | 0.0625 |
| CAR increase | 1 | 31 | 5.62 | 0.0241 |
| SSKJ 3-8 R stress coping strategies: seeking social support | 1 | 31 | 2.63 | 0.1148 |
| Time:group | 2 | 64 | 1.32 | 0.2736 |

**Table 14: Inferential statistics for spatial working memory reaction time in the ITT population, outcome was log transformed**

| | lme model summary | | | |
|---|---|---|---|---|
| | **Estimate** | **t-value** | **DF** | **p-value** |
| Intercept (placebo, baseline) | 6.622 | 68.02 | 63 | 0 |
| 1hr post dose | -0.006 | -0.17 | 63 | 0.868 |
| 2.5hr post dose | -0.019 | -0.51 | 63 | 0.609 |
| 150 mg group | 0.021 | 0.41 | 31 | 0.683 |
| Fruit and vegetable intake | -0.017 | -1.48 | 31 | 0.148 |
| Sweets intake | 0.027 | 2.41 | 31 | 0.022 |
| SSKJ 3-8 R stress coping strategies: anger related emotion regulation | 0.009 | 1.56 | 31 | 0.128 |
| SSKJ 3-8 R stress coping strategies: seeking social support | -0.012 | -2.12 | 31 | 0.042 |
| 1hr post dose:150 mg | -0.042 | -0.8 | 63 | 0.428 |
| 2.5hr post dose:150 mg | -0.037 | -0.71 | 63 | 0.479 |

| | F-tests | | | |
|---|---|---|---|---|
| | **Numerator DF** | **Denominator DF** | **F-value** | **p-value** |
| Intercept (placebo, baseline) | 1 | 63 | 98651.49 | <.001 |
| Time | 2 | 63 | 1.41 | 0.252 |
| Group | 1 | 31 | 0.17 | 0.686 |
| Fruit and vegetable intake | 1 | 31 | 6.84 | 0.014 |
| Sweets intake | | 31 | 5.82 | 0.022 |
| SSKJ 3-8 R stress coping strategies: anger related emotion regulation | | 31 | 2.36 | 0.135 |
| SSKJ 3-8 R stress coping strategies: seeking social support | | 31 | 4.56 | 0.041 |
| Time:group | 2 | 63 | 0.38 | 0.683 |

**Table 15: Inferential statistics for simple reaction time in the ITT population**

| | lme model summary | | | |
|---|---|---|---|---|
| | **Estimate** | **t-value** | **DF** | **p-value** |
| Intercept (placebo, baseline) | 327.91 | 26.19 | 66 | 0 |
| 1hr post dose | 0.623 | 0.09 | 66 | 0.927 |
| 2.5hr post dose | 8.882 | 1.32 | 66 | 0.192 |
| 150 mg group | -4.106 | -0.38 | 32 | 0.704 |
| Sweets intake | 4.224 | 1.66 | 32 | 0.106 |
| SSKJ 3-8 R Psychological stress symptoms sadness | 6.113 | 2.2 | 32 | 0.035 |
| SSKJ 3-8 R Psychological stress symptoms anxiety | 0.439 | 0.2 | 32 | 0.843 |
| 1hr post dose:150 mg | 8.605 | 0.91 | 66 | 0.369 |
| 2.5hr post dose:150 mg | 0.17 | 0.02 | 66 | 0.986 |
| | **F-tests** | | | |
| | **Numerator DF** | **Denominator DF** | **F-value** | **p-value** |
| Intercept (placebo, baseline) | 1 | 66 | 6237.228 | <.001 |
| Time | 2 | 66 | 1.966 | 0.148 |
| Group | 1 | 32 | 0.443 | 0.511 |
| Sweets intake | 1 | 32 | 5.855 | 0.021 |
| SSKJ 3-8 R Psychological stress symptoms sadness | 1 | 32 | 5.826 | 0.022 |
| SSKJ 3-8 R Psychological stress symptoms anxiety | 1 | 32 | 0.041 | 0.841 |
| Time:group | 2 | 66 | 0.548 | 0.581 |

**Table 16: Inferential statistics for digit vigilance accuracy in the ITT population**

| | lme model summary | | | |
|---|---|---|---|---|
| | **Estimate** | **t-value** | **DF** | **p-value** |
| Intercept (placebo, baseline) | 94.321 | 92.72 | 63 | <.001 |
| 1hr post dose | -0.092 | -0.08 | 63 | 0.936 |
| 2.5hr post dose | 0.778 | 0.66 | 63 | 0.511 |
| 150 mg group | 1.657 | 1.16 | 35 | 0.256 |
| 1hr post dose:150 mg | -1.496 | -0.92 | 63 | 0.361 |
| 2.5hr post dose:150 mg | -2.793 | -1.69 | 63 | 0.097 |
| | **F-tests** | | | |
| | **Numerator DF** | **Denominator DF** | **F-value** | **p-value** |
| Intercept (placebo, baseline) | 1 | 63 | 29405.99 | <.001 |
| Time | 2 | 63 | 0.57 | 0.571 |
| Group | 1 | 35 | 0.07 | 0.799 |
| Time:group | 2 | 63 | 1.43 | 0.247 |

**Table 17: Inferential statistics for digit vigilance reaction time in the ITT population**

| | lme model summary | | | |
|---|---|---|---|---|
| | **Estimate** | **t-value** | **DF** | **p-value** |
| Intercept (placebo, baseline) | 582.543 | 23.15 | 68 | 0 |
| 1hr post dose | -2.954 | -0.34 | 68 | 0.733 |
| 2.5hr post dose | 7.602 | 0.88 | 68 | 0.381 |
| 150 mg group | -11.047 | -0.94 | 33 | 0.352 |
| Sleep quality | -7.305 | -2.38 | 33 | 0.023 |
| Gender | -15.885 | -1.52 | 33 | 0.139 |
| 1hr post dose:150 mg | 6.58 | 0.54 | 68 | 0.59 |
| 2.5hr post dose:150 mg | -11.348 | -0.93 | 68 | 0.353 |
| | **F-tests** | | | |
| | **Numerator DF** | **Denominator DF** | **F-value** | **p-value** |
| Intercept (placebo, baseline) | 1 | 68 | 11614.02 | <.001 |
| Time | 2 | 68 | 0.06 | 0.944 |
| Group | 1 | 33 | 0.96 | 0.336 |
| Sleep quality | 1 | 33 | 7.48 | 0.010 |
| Gender | 1 | 33 | 2.31 | 0.138 |
| Time:group | 2 | 68 | 1.11 | 0.336 |

**Table 18: Inferential statistics for digit vigilance false alarms in the ITT population, outcome was log transformed after adding 1 to each value**

| | lme model summary | | | |
|---|---|---|---|---|
| | **Estimate** | **t-value** | **DF** | **p-value** |
| Intercept (placebo, baseline) | 0.676 | 4.62 | 64 | <.001 |
| 1hr post dose | 0.128 | 0.79 | 64 | 0.434 |
| 2.5hr post dose | 0.0003 | 0.001 | 64 | 0.999 |
| 150 mg group | 0.069 | 0.34 | 35 | 0.736 |
| 1hr post dose:150 mg | 0.064 | 0.28 | 64 | 0.777 |
| 2.5hr post dose:150 mg | 0.053 | 0.23 | 64 | 0.818 |
| | **F-tests** | | | |
| | **Numerator DF** | **Denominator DF** | **F-value** | **p-value** |
| Intercept (placebo, baseline) | 1 | 64 | 98.33 | <.001 |
| Time | 2 | 64 | 1.17 | 0.318 |
| Group | 1 | 35 | 0.47 | 0.497 |
| Time:group | 2 | 64 | 0.05 | 0.955 |

**Table 19: Inferential statistics for choice reaction time accuracy in the ITT population**

| | lme model summary | | | |
|---|---|---|---|---|
| | **Estimate** | **t-value** | **DF** | **p-value** |
| Intercept (placebo, baseline) | 102.976 | 13.65 | 62 | <.001 |
| 1hr post dose | 1.667 | 2.22 | 62 | 0.03 |
| 2.5hr post dose | 1.889 | 2.51 | 62 | 0.015 |
| 150 mg group | -0.168 | -0.16 | 33 | 0.87 |
| WHR | -14.59 | -1.64 | 33 | 0.111 |
| PDS | 0.735 | 1.49 | 33 | 0.147 |
| 1hr post dose:150 mg | 0.408 | 0.37 | 62 | 0.711 |
| 2.5hr post dose:150 mg | -0.695 | -0.63 | 62 | 0.529 |
| | **F-tests** | | | |
| | **Numerator DF** | **Denominator DF** | **F-value** | **p-value** |
| Intercept (placebo, baseline) | 1 | 62 | 51623.73 | <.001 |
| Time | 2 | 62 | 6.85 | 0.002 |
| Group | 1 | 33 | 0.10 | 0.755 |
| WHR | 1 | 33 | 3.38 | 0.075 |
| PDS | 1 | 33 | 2.25 | 0.143 |
| Time:group | 2 | 62 | 0.50 | 0.612 |
| | **Post-hoc Tukey tests for time** | | | |
| | **Estimate** | **z-value** | | **p-value** |
| 1hr post dose - baseline | 1.8098 | 3.333 | | 0.003 |
| 2.5hr post dose – baseline | 1.5441 | 2.845 | | 0.012 |
| 2.5hr post dose - 1hr post dose | -0.2658 | -0.480 | | 0.881 |

**Table 20: Inferential statistics for choice reaction time in the ITT population, outcome was log transformed**

| | lme model summary | | | |
|---|---|---|---|---|
| | **Estimate** | **t-value** | **DF** | **p-value** |
| Intercept (placebo, baseline) | 6.233 | 91.25 | 66 | 0 |
| 1hr post dose | 0.025 | 1.47 | 66 | 0.145 |
| 2.5hr post dose | 0.039 | 2.27 | 66 | 0.026 |
| 150 mg group | 0.037 | 1.13 | 32 | 0.268 |
| Fruit and vegetable intake | -0.016 | -1.93 | 32 | 0.062 |
| Sweets intake | 0.02 | 2.6 | 32 | 0.014 |
| PDS | -0.033 | -1.82 | 32 | 0.078 |
| 1hr post dose:150 mg | -0.028 | -1.15 | 66 | 0.253 |
| 2.5hr post dose:150 mg | -0.04 | -1.65 | 66 | 0.105 |
| | **F-tests** | | | |
| | **Numerator DF** | **Denominator DF** | **F-value** | **p-value** |
| Intercept (placebo, baseline) | 1 | 66 | 175419.5 | <.001 |
| Time | 2 | 66 | 1.37 | 0.261 |
| Group | 1 | 32 | 0.07 | 0.790 |
| Fruit and vegetable intake | 1 | 32 | 6.77 | 0.014 |
| Sweets intake | 1 | 32 | 5.48 | 0.026 |
| PDS | 1 | 32 | 3.21 | 0.083 |
| Time:group | 2 | 66 | 1.41 | 0.250 |

##### 3.3 Physiological outcomes

**Table 21: Inferential statistics for HR in the ITT population, outcome was log transformed**

| | lme model summary | | | |
|---|---|---|---|---|
| | **Estimate** | **t-value** | **DF** | **p-value** |
| Intercept (placebo, baseline) | 4.323 | 29.31 | 177 | <.001 |
| time | 0.0001 | 1.45 | 177 | 0.150 |
| 150 mg group | 0.036 | 0.77 | 33 | 0.446 |
| Level of activity | -0.041 | -2.87 | 33 | 0.007 |
| SSKJ 3-8 R stress vulnerability | 0.01 | 1.25 | 33 | 0.221 |
| time:150 mg | -0.0002 | -1.69 | 177 | 0.092 |
| | **F-tests** | | | |
| | **Numerator DF** | **Denominator DF** | **F-value** | **p-value** |
| Intercept (placebo, baseline) | 1 | 177 | 37387.98 | <.001 |
| Time | 1 | 177 | 0.16 | 0.694 |
| Group | 1 | 33 | 0.00 | 0.974 |
| Level of activity | 1 | 33 | 14.32 | <.001 |
| SSKJ 3-8 R stress vulnerability | 1 | 33 | 1.61 | 0.213 |
| Time:group | 1 | 177 | 2.87 | 0.092 |

Table 22: Inferential statistics for diastolic BP in the ITT population, outcome was log transformed

| | lme model summary | | | |
|---|---|---|---|---|
| | **Estimate** | **t-value** | **DF** | **p-value** |
| Intercept (placebo, baseline) | 4.02 | 28.25 | 177 | <.001 |
| time | <.001 | 0.46 | 177 | 0.645 |
| 150 mg group | -0.017 | -0.51 | 29 | 0.617 |
| Sleep disruption | 0.1 | 2.88 | 29 | 0.007 |
| PDS | 0.015 | 0.7 | 29 | 0.489 |
| Weight | 0.004 | 2.48 | 29 | 0.019 |
| Level of activity | -0.021 | -1.94 | 29 | 0.062 |
| SSKJ 3-8 R stress vulnerability | <.001 | 0.01 | 29 | 0.995 |
| SSKJ 3-8 R stress coping strategy: avoidant coping | -0.006 | -1.54 | 29 | 0.135 |
| time:150 mg | <.001 | 0.23 | 177 | 0.821 |
| | **F-tests** | | | |
| | **Numerator DF** | **Denominator DF** | **F-value** | **p-value** |
| Intercept (placebo, baseline) | 1 | 177 | 81518.24 | <.001 |
| Time | 1 | 177 | 0.95 | 0.331 |
| Group | 1 | 29 | 0.00 | 0.971 |
| Sleep disruption | 1 | 29 | 10.27 | 0.003 |
| PDS | 1 | 29 | 9.73 | 0.004 |
| Weight | 1 | 29 | 4.66 | 0.039 |
| Level of activity | 1 | 29 | 4.28 | 0.048 |
| SSKJ 3-8 R stress vulnerability | 1 | 29 | 0.01 | 0.938 |
| SSKJ 3-8 R stress coping strategy: avoidant coping | 1 | 29 | 2.39 | 0.133 |
| Time:Group | 1 | 177 | 0.05 | 0.821 |

Table 23: Inferential statistics for systolic BP in the ITT population, outcome was log transformed

| | lme model summary | | | |
|---|---|---|---|---|
| | **Estimate** | **t-value** | **DF** | **p-value** |
| Intercept (placebo, baseline) | 4.661 | 33.67 | 178 | <.001 |
| Time | <.001 | 2.8 | 178 | 0.006 |
| 150 mg group | -0.016 | -0.71 | 30 | 0.481 |
| Sleep disruption | 0.06 | 2.48 | 30 | 0.019 |
| PDS | 0.02 | 1.49 | 30 | 0.147 |
| Weight | 0.003 | 2.34 | 30 | 0.026 |
| Sweets intake | -0.011 | -2.01 | 30 | 0.054 |
| SSKJ 3-8 R Psychological stress symptoms wellbeing | -0.023 | -1.7 | 30 | 0.1 |
| time:150 mg | <.001 | -1.05 | 178 | 0.296 |
| | **lme model summary without interaction** | | | |
| | **Estimate** | **t-value** | **DF** | **p-value** |
| Intercept (placebo, baseline) | 4.664 | 33.64 | 179 | <.001 |
| Time | <.001 | 2.92 | 179 | 0.004 |
| 150 mg group | -0.023 | -1.06 | 30 | 0.299 |
| Sleep disruption | 0.06 | 2.47 | 30 | 0.019 |
| PDS | 0.02 | 1.49 | 30 | 0.148 |
| Weight | 0.003 | 2.34 | 30 | 0.026 |
| Sweets intake | -0.011 | -2 | 30 | 0.055 |
| SSKJ 3-8 R Psychological stress symptoms wellbeing | -0.023 | -1.69 | 30 | 0.101 |
| | **F-tests** | | | |
| | **Numerator DF** | **Denominator DF** | **F-value** | **p-value** |
| Intercept (placebo, baseline) | 1 | 178 | 209345.19 | <.001 |
| Time | 1 | 178 | 8.67 | 0.004 |
| Group | 1 | 30 | 0.98 | 0.331 |
| Sleep disruption | 1 | 30 | 8.04 | 0.008 |
| PDS | 1 | 30 | 6.01 | 0.020 |
| Weight | 1 | 30 | 11.13 | 0.002 |
| Sweets intake | 1 | 30 | 3.16 | 0.086 |
| SSKJ 3-8 R Psychological stress symptoms wellbeing | 1 | 30 | 2.87 | 0.100 |
| Time:Group | 1 | 178 | 1.10 | 0.296 |

**Table 24: Inferential statistics for salivary cortisol in the ITT population, outcome was square root transformed**

| | lme model summary | | | |
|---|---|---|---|---|
| | **Estimate** | **t-value** | **DF** | **p-value** |
| Intercept (placebo, baseline) | 1.819 | 4.38 | 173 | <.001 |
| Time | 0.001 | 2.09 | 173 | 0.038 |
| 150 mg group | 0.039 | 0.49 | 28 | 0.631 |
| Gender | 0.157 | 2.02 | 28 | 0.053 |
| Sleep duration | -0.045 | -2.72 | 28 | 0.011 |
| Sweets intake | 0.019 | 1.06 | 28 | 0.300 |
| Level of activity | -0.06 | -2.08 | 28 | 0.047 |
| SSKJ 3-8 R Physical stress symptoms | 0.024 | 0.97 | 28 | 0.338 |
| SSKJ 3-8 R stress vulnerability | 0.013 | 0.93 | 28 | 0.361 |
| Time:150 mg | -0.001 | -1.51 | 173 | 0.133 |

| | F-tests | | | |
|---|---|---|---|---|
| | **Numerator DF** | **Denominator DF** | **F-value** | **p-value** |
| Intercept (placebo, baseline) | 1 | 173 | 2355.43 | <.001 |
| Time | 1 | 173 | 1.49 | 0.224 |
| Group | 1 | 28 | 0.20 | 0.655 |
| Gender | 1 | 28 | 10.87 | 0.003 |
| Sleep duration | 1 | 28 | 10.63 | 0.003 |
| Sweets intake | 1 | 28 | 4.18 | 0.051 |
| Level of activity | 1 | 28 | 8.59 | 0.007 |
| SSKJ 3-8 R Physical stress symptoms | 1 | 28 | 1.00 | 0.326 |
| SSKJ 3-8 R stress vulnerability | 1 | 28 | 0.50 | 0.485 |
| Time:Group | 1 | 173 | 2.27 | 0.134 |

Table 25: Inferential statistics for salivary oxytocin in the ITT population

| | lme model summary | | | |
|---|---|---|---|---|
| | **Estimate** | **t-value** | **DF** | **p-value** |
| Intercept (placebo, before treatment) | 1.190 | 7.19 | 34 | <.001 |
| time (after treatment) | -0.176 | -2.12 | 34 | 0.042 |
| 150 mg group | 0.048 | 0.37 | 32 | 0.712 |
| SSKJ 3-8 R stress coping strategy: avoidant coping | 0.031 | 1.83 | 32 | 0.077 |
| SSKJ 3-8 R stress coping strategy: palliative emotion regulation | 0.024 | 1.42 | 32 | 0.166 |
| time:150 mg | 0.030 | 0.26 | 34 | 0.800 |
| | | | **lme model summary without interaction** | |
| | **Estimate** | **t-value** | **DF** | **p-value** |
| Intercept (placebo, before treatment) | 1.182 | 7.26 | 35 | <.001 |
| time (after treatment) | -0.161 | -2.78 | 35 | 0.009 |
| 150 mg group | 0.063 | 0.55 | 32 | 0.588 |
| SSKJ 3-8 R stress coping strategy: avoidant coping | 0.031 | 1.83 | 32 | 0.077 |
| SSKJ 3-8 R stress coping strategy: palliative emotion regulation | 0.024 | 1.42 | 32 | 0.166 |
| | | **F-tests** | | |
| | **Numerator DF** | **Denominator DF** | **F-value** | **p-value** |
| Intercept (placebo, before treatment) | 1 | 34 | 778.2795 | <.001 |
| Time | 1 | 34 | 7.5052 | 0.010 |
| Group | 1 | 32 | 0.4414 | 0.511 |
| SSKJ 3-8 R stress coping strategy: avoidant coping | 1 | 32 | 7.8666 | 0.009 |
| SSKJ 3-8 R stress coping strategy: palliative emotion regulation | 1 | 32 | 2.0133 | 0.166 |
| Time:Group | 1 | 34 | 0.0655 | 0.800 |

#### 3.4 Exploratory cognitive outcomes

**Table 26: Inferential statistics for secondary memory in the ITT population**

| | lme model summary | | | |
|---|---|---|---|---|
| | **Estimate** | **t-value** | **DF** | **p-value** |
| Intercept (placebo, baseline) | 173.267 | 8.06 | 68 | <.001 |
| 1hr post dose | -18.102 | -3.19 | 68 | 0.002 |
| 2.5hr post dose | -13.472 | -2.38 | 68 | 0.020 |
| 150 mg group | -2.674 | -0.29 | 33 | 0.776 |
| Sleep quality | 4.321 | 1.63 | 33 | 0.112 |
| Gender | 26.472 | 2.94 | 33 | 0.006 |
| 1hr post dose:150 mg | 0.077 | 0.01 | 68 | 0.992 |
| 2.5hr post dose:150 mg | -9.072 | -1.13 | 68 | 0.261 |
| | **F-tests** | | | |
| | **Numerator DF** | **Denominator DF** | **F-value** | **p-value** |
| Intercept (placebo, baseline) | 1 | 68 | 3109.13 | <.001 |
| Time | 2 | 68 | 13.44 | <.001 |
| Group | 1 | 33 | 1.19 | 0.284 |
| Sleep quality | 1 | 33 | 5.12 | 0.030 |
| Gender | 1 | 33 | 8.59 | 0.006 |
| Time:group | 2 | 68 | 0.86 | 0.427 |
| | **Post-hoc Tukey tests for time** | | | |
| | **Estimate** | **z-value** | | **p-value** |
| 1hr post dose - baseline | -18.078 | -4.528 | | < 0.001 |
| 2.5hr post dose – baseline | -18.023 | -4.514 | | < 0.001 |
| 2.5hr post dose - 1hr post dose | 0.055 | 0.014 | | 1 |

**Table 27: Inferential statistics for working memory in the ITT population, outcome was square transformed**

| | lme model summary | | | |
|---|---|---|---|---|
| | **Estimate** | **t-value** | **DF** | **p-value** |
| Intercept (placebo, baseline) | 34416.9 | 28.34 | 64 | <.001 |
| 1hr post dose | 2409.693 | 2.89 | 64 | 0.005 |
| 2.5hr post dose | 1078.681 | 1.34 | 64 | 0.184 |
| 150 mg group | 2338.597 | 2.70 | 32 | 0.011 |
| BFP | -146.788 | -3.44 | 32 | 0.002 |
| SSKJ 3-8 R<br>stress coping strategies:<br>seeking social support | 210.920 | 2.49 | 32 | 0.018 |
| SSKJ 3-8 R<br>Psychological stress symptoms<br>sadness | 54.783 | 0.28 | 32 | 0.778 |
| 1hr post dose:150 mg | -2527.030 | -2.21 | 64 | 0.03 |
| 2.5hr post dose:150 mg | -1255.140 | -1.12 | 64 | 0.266 |

| | F-tests | | | |
|---|---|---|---|---|
| | **Numerator DF** | **Denominator DF** | **F-value** | **p-value** |
| Intercept (placebo, baseline) | 1 | 64 | 15141.08 | <.001 |
| Time | 2 | 64 | 1.97 | 0.148 |
| Group | 1 | 32 | 2.24 | 0.144 |
| BFP | 1 | 32 | 10.41 | 0.003 |
| SSKJ 3-8 R<br>stress coping strategies:<br>seeking social support | 1 | 32 | 7.09 | 0.012 |
| SSKJ 3-8 R<br>Psychological stress symptoms<br>sadness | 1 | 32 | 0.06 | 0.807 |
| Time:group | 2 | 64 | 2.46 | 0.094 |

**Table 28: Inferential statistics for speed of memory in the ITT population, outcome was log transformed**

| | **lme model summary** | | | |
|---|---|---|---|---|
| | **Estimate** | **t-value** | **DF** | **p-value** |
| Intercept (placebo, baseline) | 8.441 | 57.98 | 66 | <.001 |
| 1hr post dose | -0.02 | -0.89 | 66 | 0.376 |
| 2.5hr post dose | -0.068 | -3.00 | 66 | 0.004 |
| 150 mg group | -0.007 | -0.15 | 32 | 0.879 |
| Weight | -0.004 | -1.83 | 32 | 0.077 |
| Fruit and vegetable intake | -0.022 | -2.03 | 32 | 0.051 |
| Sweets intake | 0.021 | 1.92 | 32 | 0.064 |
| 1hr post dose:150 mg | 0.03 | 0.96 | 66 | 0.343 |
| 2.5hr post dose:150 mg | 0.052 | 1.63 | 66 | 0.108 |
| | **F-tests** | | | |
| | **Numerator DF** | **Denominator DF** | **F-value** | **p-value** |
| Intercept (placebo, baseline) | 1 | 66 | 160852.5 | <.001 |
| Time | 2 | 66 | 4.19 | 0.019 |
| Group | 1 | 32 | 0.14 | 0.716 |
| Weight | 1 | 32 | 4.08 | 0.052 |
| Fruit and vegetable intake | 1 | 32 | 4.94 | 0.033 |
| Sweets intake | 1 | 32 | 3.62 | 0.066 |
| Time:group | 2 | 66 | 1.34 | 0.269 |
| | **Post-hoc Tukey tests for time** | | | |
| | **Estimate** | **z-value** | | **p-value** |
| 1hr post dose - baseline | -0.005 | -0.301 | | 0.951 |
| 2.5hr post dose – baseline | -0.042 | -2.599 | | 0.025 |
| 2.5hr post dose - 1hr post dose | -0.037 | -2.301 | | 0.056 |

**Table 29: Inferential statistics for accuracy of attention in the ITT population**

| | lme model summary | | | |
|---|---|---|---|---|
| | **Estimate** | **t-value** | **DF** | **p-value** |
| Intercept (placebo, baseline) | 93.883 | 111.26 | 68 | <.001 |
| 1hr post dose | 0.402 | 0.44 | 68 | 0.662 |
| 2.5hr post dose | 0.512 | 0.56 | 68 | 0.577 |
| 150 mg group | 0.41 | 0.35 | 35 | 0.73 |
| 1hr post dose:150 mg | -0.588 | -0.46 | 68 | 0.65 |
| 2.5hr post dose:150 mg | -1.983 | -1.54 | 68 | 0.129 |
| | **F-tests** | | | |
| | **Numerator DF** | **Denominator DF** | **F-value** | **p-value** |
| Intercept (placebo, baseline) | 1 | 68 | 41493.89 | <.001 |
| Time | 2 | 68 | 0.47 | 0.628 |
| Group | 1 | 35 | 0.22 | 0.642 |
| Time:group | 2 | 68 | 1.24 | 0.294 |

**Table 30: Inferential statistics for speed of attention in the ITT population**

| | lme model summary | | | |
|---|---|---|---|---|
| | **Estimate** | **t-value** | **DF** | **p-value** |
| Intercept (placebo, baseline) | 1472.81 | 14.47 | 67 | <.001 |
| 1hr post dose | 8.34 | 0.54 | 67 | 0.593 |
| 2.5hr post dose | 34.496 | 2.22 | 67 | 0.030 |
| 150 mg group | 13.856 | 0.41 | 30 | 0.682 |
| PDS | -26.347 | -1.40 | 30 | 0.173 |
| Weight | -1.800 | -1.10 | 30 | 0.280 |
| Fruit and vegetable intake | -9.446 | -1.17 | 30 | 0.252 |
| Sweets intake | 15.475 | 1.93 | 30 | 0.063 |
| SSKJ 3-8 R Psychological stress symptoms sadness | 10.322 | 1.20 | 30 | 0.238 |
| 1hr post dose:150 mg | 6.587 | 0.30 | 67 | 0.767 |
| 2.5hr post dose:150 mg | -24.911 | -1.13 | 67 | 0.265 |

| | F-tests | | | |
|---|---|---|---|---|
| | **Numerator DF** | **Denominator DF** | **F-value** | **p-value** |
| Intercept (placebo, baseline) | 1 | 67 | 8912.48 | <.001 |
| Time | 2 | 67 | 2.08 | 0.133 |
| Group | 1 | 30 | 0.15 | 0.703 |
| PDS | 1 | 30 | 4.22 | 0.049 |
| Weight | 1 | 30 | 1.56 | 0.222 |
| Fruit and vegetable intake | 1 | 30 | 3.37 | 0.076 |
| Sweets intake | 1 | 30 | 5.32 | 0.028 |
| SSKJ 3-8 R Psychological stress symptoms sadness | 1 | 30 | 1.43 | 0.241 |
| Time:group | 2 | 67 | 1.14 | 0.325 |

**Table 31: Inferential statistics for Picture Recognition Original Stimuli Accuracy in the ITT population, outcome was square transformed**

| | Linear mixed effects model summary | | | |
|---|---|---|---|---|
| | **Estimate** | **t-value** | **DF** | **p-value** |
| Intercept (placebo, baseline) | 6455.795 | 9.02 | 66 | <.001 |
| 1hr post dose | -613.889 | -1.39 | 66 | 0.170 |
| 2.5hr post dose | 52.963 | 0.12 | 66 | 0.907 |
| 150 mg | -364.779 | -0.67 | 34 | 0.507 |
| Gender | 802.946 | 1.78 | 34 | 0.084 |
| 1hr post dose:150 mg | 39.998 | 0.06 | 66 | 0.949 |
| 2.5hr post dose:150 mg | -73.752 | -0.12 | 66 | 0.908 |
| | **F-tests** | | | |
| | **Numerator DF** | **Denominator DF** | **F-value** | **p-value** |
| Intercept (placebo, baseline) | 1 | 66 | 1189.1941 | <.001 |
| Time | 2 | 66 | 2.4478 | 0.094 |
| Group | 1 | 34 | 1.064 | 0.310 |
| Gender | 1 | 34 | 3.1773 | 0.084 |
| Time:group | 2 | 66 | 0.0163 | 0.984 |

**Table 32: Inferential statistics for Picture Recognition New Stimuli Accuracy in the ITT population**

| | Linear mixed effects model summary | | | |
|---|---|---|---|---|
| | **Estimate** | **t-value** | **DF** | **p-value** |
| Intercept (placebo, baseline) | 85.44456 | 18.01 | 68 | <.001 |
| 1hr post dose | -5.83333 | -1.95 | 68 | 0.056 |
| 2.5hr post dose | -5.27778 | -1.76 | 68 | 0.082 |
| 150 mg | -3.95199 | -1.09 | 33 | 0.282 |
| Sweets intake | 1.64586 | 2.36 | 33 | 0.024 |
| SSKJ 3-8 R Palliative emotion regulation | -0.71427 | -1.97 | 33 | 0.057 |
| 1hr post dose:150 mg | 0.63935 | 0.15 | 68 | 0.880 |
| 2.5hr post dose:150 mg | 1.68935 | 0.40 | 68 | 0.690 |
| | **F-tests** | | | |
| | **Numerator DF** | **Denominator DF** | **F-value** | **p-value** |
| Intercept (placebo, baseline) | 1 | 68 | 3564.589 | <.001 |
| Time | 2 | 68 | 3.977 | 0.023 |
| Group | 1 | 33 | 2.756 | 0.106 |
| Sweets intake | 1 | 33 | 4.774 | 0.036 |
| SSKJ 3-8 R Palliative emotion regulation | 1 | 33 | 3.862 | 0.058 |
| Time:group | 2 | 68 | 0.082 | 0.922 |

| | Post-hoc Tukey tests for time | | |
|---|---|---|---|
| | **Estimate** | **z-value** | **p-value** |
| 1hr post dose - baseline | -5.508 | -2.647 | 0.0219 |
| 2.5hr post dose – baseline | -4.427 | -2.128 | 0.0843 |
| 2.5hr post dose - 1hr post dose | 1.081 | 0.517 | 0.8629 |
