## Supplementary Data S4 for "Assessment of the Effects of a Sage (*Salvia officinalis*) Extract on Cognitive Performance in Adolescents and Young Adults"

### 4 Supplementary Data File S4

#### 4.1 Baseline characteristics and control variables

**Table 1: Inferential statistics for baseline characteristics and control variables**

| F-tests for continuous control variables | | | | |
|---|---|---|---|---|
| Control variable | **Numerator DF** | **Denominator DF** | **F-value** | **p-value** |
| TICS chronical stress screening scale | 2 | 33 | 2.895 | 0.069 |
| CAR increase | 2 | 33 | 1.418 | 0.257 |
| Sleep quality | 2 | 33 | 0.472 | 0.628 |
| Sleep duration | 2 | 33 | 0.570 | 0.571 |
| Fruit and vegetable intake | 2 | 33 | 0.147 | 0.864 |
| Sweets intake | 2 | 33 | 0.179 | 0.837 |
| Level of activity | 2 | 33 | 0.282 | 0.756 |
| Weight | 2 | 33 | 0.055 | 0.947 |
| BMI | 2 | 33 | 0.236 | 0.791 |
| WHR | 2 | 33 | 0.365 | 0.697 |
| BFP | 2 | 33 | 0.722 | 0.493 |
| MP | 2 | 33 | 0.658 | 0.525 |
| **Fisher's exact test for categorical control variables** | | | | |
| | | | | **p-value** |
| Gender | | | | 0.903 |
| **Post-hoc Welch t-tests for TICS SCCS** | | | | |
| | | **DF** | **t-value** | **p-value** |
| Placebo vs 150 mg group | | **18.51** | 0.12 | 0.904 |
| Placebo vs 300 mg group | | 17.65 | -2.12 | 0.049 |

#### 4.2 Cognitive outcomes

**Table 2: Inferential statistics for Picture Recognition Accuracy in the ITT population, outcome was square transformed**

| | Linear mixed effects model summary | | | |
|---|---|---|---|---|
| | **Estimate** | **t-value** | **DF** | **p-value** |
| Intercept (placebo, baseline) | 4714.485 | 4.86 | 64 | <.001 |
| 1hr post dose | -99.279 | -0.22 | 64 | 0.823 |
| 2.5hr post dose | 319.992 | 0.72 | 64 | 0.472 |
| 150 mg group | 383.811 | 0.61 | 29 | 0.545 |
| 300 mg group | 358.749 | 0.55 | 29 | 0.589 |
| CAR increase | 58.502 | 1.96 | 29 | 0.060 |
| BFP | 16.279 | 0.59 | 29 | 0.557 |
| TICS social overload | 101.760 | 1.93 | 29 | 0.063 |
| Gender | 360.789 | 0.70 | 29 | 0.492 |
| 1hr post dose:150 mg | -237.179 | -0.38 | 64 | 0.702 |
| 2.5hr post dose:150 mg | -868.950 | -1.41 | 64 | 0.163 |
| 1hr post dose:300 mg | 174.458 | 0.28 | 64 | 0.781 |
| 2.5hr post dose:300 mg | -115.646 | -0.19 | 64 | 0.854 |
| | **F-tests** | | | |
| | **Numerator DF** | **Denominator DF** | **F-value** | **p-value** |
| Intercept (placebo, baseline) | 1 | 64 | 1314.48 | <.001 |
| Time | 2 | 64 | 0.20 | 0.817 |
| Group | 2 | 29 | 0.48 | 0.624 |
| CAR increase | 1 | 29 | 4.41 | 0.044 |
| BFP | 1 | 29 | 2.63 | 0.116 |
| TICS social overload | 1 | 29 | 3.96 | 0.056 |
| Gender | 1 | 29 | 0.48 | 0.493 |
| Time:group | 4 | 64 | 0.65 | 0.628 |

**Table 3: Inferential statistics for Picture Recognition Accuracy in the PP population**

| | Linear mixed effects model summary | | | |
|---|---|---|---|---|
| | **Estimate** | **t-value** | **DF** | **p-value** |
| Intercept (placebo, baseline) | 68.499 | 11.61 | 60 | <.001 |
| 1hr post dose | -1.111 | -0.39 | 60 | 0.696 |
| 2.5hr post dose | 0.833 | 0.29 | 60 | 0.770 |
| 150 mg group | 2.827 | 0.67 | 27 | 0.506 |
| 300 mg group | 1.377 | 0.32 | 27 | 0.749 |
| CAR increase | 0.188 | 1.05 | 27 | 0.301 |
| BFP | 0.499 | 2.57 | 27 | 0.016 |
| TICS social overload | 0.650 | 1.88 | 27 | 0.071 |
| 1hr post dose:150 mg | -0.972 | -0.26 | 60 | 0.796 |
| 2.5hr post dose:150 mg | -4.167 | -1.11 | 60 | 0.271 |
| 1hr post dose:300 mg | 2.778 | 0.74 | 60 | 0.462 |
| 2.5hr post dose:300 mg | 1.667 | 0.44 | 60 | 0.658 |
| | **F-tests** | | | |
| | **Numerator DF** | **Denominator DF** | **F-value** | **p-value** |
| Intercept (placebo, baseline) | 1 | 60 | 4084.814 | <.001 |
| Time | 2 | 60 | 0.054 | 0.947 |
| Group | 2 | 27 | 0.375 | 0.691 |
| CAR increase | 1 | 27 | 4.630 | 0.041 |
| BFP | 1 | 27 | 6.217 | 0.019 |
| TICS social overload | 1 | 27 | 3.531 | 0.071 |
| Time:group | 4 | 60 | 0.832 | 0.510 |

**Table 4: Inferential statistics for Picture Recognition Reaction Time in the ITT population**

| | Linear mixed effects model summary | | | |
|---|---|---|---|---|
| | **Estimate** | **t-value** | **DF** | **p-value** |
| Intercept (placebo, baseline) | 1166.708 | 17.01 | 63 | <.001 |
| 1hr post dose | -29.812 | -0.96 | 63 | 0.341 |
| 2.5hr post dose | -68.748 | -2.15 | 63 | 0.036 |
| 150 mg group | 80.334 | 1.50 | 29 | 0.144 |
| 300 mg group | 18.374 | 0.32 | 29 | 0.750 |
| TICS chronic stress | 5.318 | 2.47 | 29 | 0.020 |
| Level of activity | -33.875 | -3.27 | 29 | 0.003 |
| Sleep disruptions | -117.785 | -2.75 | 29 | 0.010 |
| 1hr post dose:150 mg | -51.673 | -1.18 | 63 | 0.244 |
| 2.5hr post dose:150 mg | -36.035 | -0.81 | 63 | 0.422 |
| 1hr post dose:300 mg | -5.716 | -0.13 | 63 | 0.899 |
| 2.5hr post dose:300 mg | 17.903 | 0.39 | 63 | 0.696 |

| | F-tests | | | |
|---|---|---|---|---|
| | **Numerator DF** | **Denominator DF** | **F-value** | **p-value** |
| Intercept (placebo, baseline) | 1 | 63 | 2861.283 | <.001 |
| Time | 2 | 63 | 8.889 | 0.004 |
| Group | 2 | 29 | 0.204 | 0.817 |
| TICS chronic stress | 1 | 29 | 7.211 | 0.012 |
| Level of activity | 1 | 29 | 7.882 | 0.009 |
| Sleep disruptions | 1 | 29 | 7.564 | 0.010 |
| Time:group | 4 | 63 | 0.575 | 0.682 |

| | Post-hoc Tukey tests for time | | |
|---|---|---|---|
| | **Estimate** | **z-value** | **p-value** |
| 1hr post dose - baseline | -49.32 | -2.746 | 0.017 |
| 2.5hr post dose – baseline | -75.68 | -4.169 | <0.001 |
| 2.5hr post dose - 1hr post dose | -26.35 | -1.452 | 0.314 |

**Table 5: Inferential statistics for Word Recognition Accuracy in the ITT population**

| | Linear mixed effects model summary | | | |
|---|---|---|---|---|
| | **Estimate** | **t-value** | **DF** | **p-value** |
| Intercept (placebo, baseline) | 97.101 | 15.60 | 65 | <.001 |
| 1hr post dose | -2.768 | -0.91 | 65 | 0.365 |
| 2.5hr post dose | -5.219 | -1.77 | 65 | 0.081 |
| 150 mg group | -3.66 | -0.93 | 32 | 0.360 |
| 300 mg group | 0.514 | 0.13 | 32 | 0.897 |
| Weight | -0.163 | -2.11 | 32 | 0.043 |
| 1hr post dose:150 mg | 1.082 | 0.26 | 65 | 0.799 |
| 2.5hr post dose:150 mg | -1.329 | -0.32 | 65 | 0.750 |
| 1hr post dose:300 mg | 1.1400 | 0.27 | 65 | 0.788 |
| 2.5hr post dose:300 mg | 3.314 | 0.80 | 65 | 0.429 |
| | **F-tests** | | | |
| | **Numerator DF** | **Denominator DF** | **F-value** | **p-value** |
| Intercept (placebo, baseline) | 1 | 65 | 4155.099 | <.001 |
| Time | 2 | 65 | 3.627 | 0.032 |
| Group | 2 | 32 | 1.603 | 0.217 |
| Weight | 1 | 32 | 4.410 | 0.044 |
| Time:group | 4 | 65 | 0.438 | 0.781 |
| | **Post-hoc Tukey tests for time** | | | |
| | **Estimate** | **z-value** | | **p-value** |
| 1hr post dose - baseline | -2.019 | -1.196 | | 0.455 |
| 2.5hr post dose – baseline | -4.557 | -2.727 | | 0.018 |
| 2.5hr post dose - 1hr post dose | -2.538 | -1.504 | | 0.289 |

**Table 6: Inferential statistics for Word Recognition Reaction Time in the ITT population**

| | Linear mixed effects model summary | | | |
|---|---|---|---|---|
| | **Estimate** | **t-value** | **DF** | **p-value** |
| Intercept (placebo, baseline) | 632.365 | 7 | 65 | <.0001 |
| 1hr post dose | -1.263 | -0.04 | 65 | 0.965 |
| 2.5hr post dose | 14.047 | 0.48 | 65 | 0.631 |
| 150 mg group | 25.945 | 0.73 | 29 | 0.473 |
| 300 mg group | -1.325 | -0.04 | 29 | 0.971 |
| Sleep disruptions | -85.135 | -3.69 | 29 | 0.001 |
| Sleep quality | 18.649 | 2.33 | 29 | 0.027 |
| Sweets intake | 0.604 | 0.09 | 29 | 0.925 |
| TICS pressure to succeed | 5.733 | 2.8 | 29 | 0.009 |
| 1hr post dose:150 mg | 13.402 | 0.32 | 65 | 0.749 |
| 2.5hr post dose:150 mg | -26.49 | -0.64 | 65 | 0.528 |
| 1hr post dose:300 mg | 38.814 | 0.94 | 65 | 0.349 |
| 2.5hr post dose:300 mg | 38.023 | 0.92 | 65 | 0.359 |
| | **F-tests** | | | |
| | **Numerator DF** | **Denominator DF** | **F-value** | **p-value** |
| Intercept (placebo, baseline) | 1 | 65 | 6089.959 | <.001 |
| Time | 2 | 65 | 0.691 | 0.505 |
| Group | 2 | 29 | 0.355 | 0.704 |
| Sleep disruptions | 1 | 29 | 17.373 | <.001 |
| Sleep quality | 1 | 29 | 5.782 | 0.023 |
| Sweets intake | 1 | 29 | 3.633 | 0.067 |
| TICS pressure to succeed | 1 | 29 | 7.824 | 0.009 |
| Time:group | 4 | 65 | 0.764 | 0.552 |

**Table 7: Inferential statistics for Immediate Word Recall Accuracy in the ITT population**

| | Linear mixed effects model summary | | | |
|---|---|---|---|---|
| | Estimate | t-value | DF | p-value |
| Intercept (placebo, baseline) | 70.025 | 5.42 | 66 | <.001 |
| 1hr post dose | -15.357 | -4.17 | 66 | <.001 |
| 2.5hr post dose | -6.189 | -1.68 | 66 | 0.098 |
| 150 mg group | -8.557 | -1.64 | 30 | 0.112 |
| 300 mg group | -1.313 | -0.24 | 30 | 0.811 |
| Weight | -0.163 | -1.46 | 30 | 0.155 |
| Gender | 6.73 | 1.63 | 30 | 0.114 |
| TICS social overload | -1.082 | -2.38 | 30 | 0.024 |
| 1hr post dose:150 mg | 14.88 | 2.85 | 66 | 0.006 |
| 2.5hr post dose:150 mg | 2.617 | 0.50 | 66 | 0.617 |
| 1hr post dose:300 mg | 10.437 | 2.00 | 66 | 0.049 |
| 2.5hr post dose:300 mg | 5.157 | 0.99 | 66 | 0.326 |
| | **F-tests** | | | |
| | **Numerator DF** | **Denominator DF** | **F-value** | **p-value** |
| Intercept (placebo, baseline) | 1 | 66 | 996.778 | <.001 |
| Time | 2 | 66 | 5.285 | 0.007 |
| Group | 2 | 30 | 0.311 | 0.735 |
| Weight | 1 | 30 | 5.122 | 0.031 |
| Gender | 1 | 30 | 1.353 | 0.254 |
| TICS social overload | 1 | 30 | 5.676 | 0.024 |
| Time:group | 4 | 66 | 2.522 | 0.049 |
| | **Post-hoc Tukey tests for time** | | | |
| | **Estimate** | **z-value** | | **p-value** |
| 1hr post dose - baseline | -6.918 | -3.117 | | 0.005 |
| 2.5hr post dose – baseline | -3.598 | -1.621 | | 0.237 |
| 2.5hr post dose - 1hr post dose | 3.320 | 1.496 | | 0.293 |

**Table 8: Inferential statistics for Immediate Word Recall Errors in the ITT population**

| | glm model summary | | | |
|---|---|---|---|---|
| | **Estimate** | **z-value** | **DF** | **p-value** |
| Intercept (placebo, baseline) | -4.810 | -3.660 | 60 | <.001 |
| 1hr post dose | 2.212 | 1.796 | 60 | 0.073 |
| 2.5hr post dose | 1.370 | 1.080 | 60 | 0.290 |
| 150 mg group | 1.551 | 1.205 | 27 | 0.228 |
| 300 mg group | 0.647 | 0.495 | 27 | 0.621 |
| TICS pressure to succeed | 0.137 | 2.129 | 27 | 0.033 |
| TICS social overload | 0.037 | 0.460 | 27 | 0.646 |
| TICS work overload | 0.016 | 0.327 | 27 | 0.744 |
| 1hr post dose:150 mg | -4.411 | -2.366 | 60 | 0.018 |
| 2.5hr post dose:150 mg | -2.558 | -1.503 | 60 | 0.133 |
| 1hr post dose:300 mg | -1.726 | -1.093 | 60 | 0.275 |
| 2.5hr post dose:300 mg | -1.370 | -0.838 | 60 | 0.402 |

| | LR-tests | | |
|---|---|---|---|
| | **DF** | **LR Chisquare-value** | **p-value** |
| Time | 2 | 0.4661 | 0.792 |
| Group | 2 | 1.2919 | 0.524 |
| TICS pressure to succeed | 1 | 5.0910 | 0.024 |
| TICS social overload | 1 | 0.2107 | 0.646 |
| TICS work overload | 1 | 0.1067 | 0.744 |
| Time:group | 4 | 7.1263 | 0.129 |

**Table 9: Inferential statistics for Delayed Word Recall Accuracy in the ITT population**

| | Linear mixed effects model summary | | | |
|---|---|---|---|---|
| | **Estimate** | **t-value** | **DF** | **p-value** |
| Intercept (placebo, baseline) | 60.722 | 8.72 | 66 | <.001 |
| 1hr post dose | -14.244 | -3.03 | 66 | 0.004 |
| 2.5hr post dose | -18.098 | -3.85 | 66 | <.001 |
| 150 mg group | -8.789 | -1.25 | 32 | 0.221 |
| 300 mg group | -3.317 | -0.46 | 32 | 0.647 |
| TICS pressure to succeed | -0.754 | -1.97 | 32 | 0.058 |
| 1hr post dose:150 mg | 7.22 | 1.08 | 66 | 0.282 |
| 2.5hr post dose:150 mg | 5.797 | 0.87 | 66 | 0.387 |
| 1hr post dose:300 mg | 2.737 | 0.41 | 66 | 0.682 |
| 2.5hr post dose:300 mg | 12.144 | 1.82 | 66 | 0.073 |
| | **F-tests** | | | |
| | **Numerator DF** | **Denominator DF** | **F-value** | **p-value** |
| Intercept (placebo, baseline) | 1 | 66 | 252.050 | <.001 |
| Time | 2 | 66 | 12.080 | <.001 |
| Group | 2 | 32 | 0.495 | 0.614 |
| TICS pressure to succeed | 1 | 32 | 3.865 | 0.058 |
| Time:group | 4 | 66 | 1.276 | 0.288 |
| | **Post-hoc Tukey tests for time** | | | |
| | **Estimate** | **z-value** | | **p-value** |
| 1hr post dose - baseline | -10.925 | -3.989 | | <.001 |
| 2.5hr post dose – baseline | -12.117 | -4.424 | | <.001 |
| 2.5hr post dose - 1hr post dose | -1.192 | -0.435 | | 0.901 |

**Table 10: Inferential statistics for Delayed Word Recall Errors in the ITT population, outcome was log transformed after adding 1 to each value**

| | Linear mixed effects model summary | | | |
|---|---|---|---|---|
| | **Estimate** | **t-value** | **DF** | **p-value** |
| Intercept (placebo, baseline) | 0.011 | 0.04 | 56 | 0.971 |
| 1hr post dose | -0.044 | -0.21 | 56 | 0.833 |
| 2.5hr post dose | 0.441 | 2.31 | 56 | 0.024 |
| 150 mg group | -0.096 | -0.43 | 28 | 0.669 |
| 300 mg group | -0.177 | -0.82 | 28 | 0.418 |
| Sweets intake | -0.002 | -0.07 | 28 | 0.948 |
| TICS work overload | 0.011 | 1.02 | 28 | 0.318 |
| TICS social overload | 0.015 | 0.73 | 28 | 0.473 |
| TICS pressure to succeed | 0.018 | 1.16 | 28 | 0.257 |
| TICS social isolation | -0.003 | -0.17 | 28 | 0.867 |
| 1hr post dose:150 mg | 0.101 | 0.35 | 56 | 0.728 |
| 2.5hr post dose:150 mg | -0.436 | -1.59 | 56 | 0.118 |
| 1hr post dose:300 mg | 0.172 | 0.6 | 56 | 0.549 |
| 2.5hr post dose:300 mg | -0.341 | -1.28 | 56 | 0.206 |
| | **F-tests** | | | |
| | **Numerator DF** | **Denominator DF** | **F-value** | **p-value** |
| Intercept (placebo, baseline) | 1 | 56 | 71.467 | <.001 |
| Time | 2 | 56 | 1.963 | 0.150 |
| Group | 2 | 28 | 1.503 | 0.240 |
| Sweets intake | 1 | 28 | 3.635 | 0.067 |
| TICS work overload | 1 | 28 | 2.834 | 0.103 |
| TICS social overload | 1 | 28 | 1.726 | 0.200 |
| TICS pressure to succeed | 1 | 28 | 1.422 | 0.243 |
| TICS social isolation | 1 | 28 | 0.007 | 0.935 |
| Time:group | 4 | 56 | 1.308 | 0.278 |

**Table 11: Inferential statistics for Numeric Working Memory Accuracy in the ITT population**

| | Linear mixed effects model summary | | | |
|---|---|---|---|---|
| | **Estimate** | **t-value** | **DF** | **p-value** |
| Intercept (placebo, baseline) | 95.399 | 50.51 | 62 | <.001 |
| 1hr post dose | -0.84 | -0.58 | 62 | 0.567 |
| 2.5hr post dose | -0.606 | -0.41 | 62 | 0.685 |
| 150 mg group | -2.585 | -1.59 | 31 | 0.122 |
| 300 mg group | 0.034 | 0.02 | 31 | 0.983 |
| Sleep disruptions | -2.122 | -2.18 | 31 | 0.037 |
| Fruit and vegetable intake | 0.422 | 1.64 | 31 | 0.111 |
| 1hr post dose:150 mg | 2.786 | 1.35 | 62 | 0.182 |
| 2.5hr post dose:150 mg | 2.086 | 1.00 | 62 | 0.321 |
| 1hr post dose:300 mg | 2.506 | 1.23 | 62 | 0.224 |
| 2.5hr post dose:300 mg | 1.161 | 0.56 | 62 | 0.574 |
| | **F-tests** | | | |
| | **Numerator DF** | **Denominator DF** | **F-value** | **p-value** |
| Intercept (placebo, baseline) | 1 | 62 | 47155.21 | <.001 |
| Time | 2 | 62 | 0.65 | 0.525 |
| Group | 2 | 31 | 2.55 | 0.094 |
| Sleep disruptions | 1 | 31 | 4.68 | 0.038 |
| Fruit and vegetable intake | 1 | 31 | 2.80 | 0.104 |
| Time:group | 4 | 62 | 0.61 | 0.654 |

**Table 12: Inferential statistics for Numeric Working Memory Reaction Time in the ITT population, outcome was log transformed**

| | Linear mixed effects model summary | | | |
|---|---|---|---|---|
| | **Estimate** | **t-value** | **DF** | **p-value** |
| Intercept (placebo, baseline) | 6.359 | 72.89 | 62 | <.001 |
| 1hr post dose | -0.023 | -0.74 | 62 | 0.462 |
| 2.5hr post dose | -0.045 | -1.47 | 62 | 0.146 |
| 150 mg group | 0.035 | 0.64 | 28 | 0.526 |
| 300 mg group | -0.019 | -0.34 | 28 | 0.735 |
| TICS chronic stress | 0.000 | -0.05 | 28 | 0.961 |
| Sweets intake | -0.003 | -0.26 | 28 | 0.794 |
| TICS social overload | -0.004 | -0.64 | 28 | 0.530 |
| TICS pressure to succeed | 0.010 | 2.14 | 28 | 0.041 |
| TICS social isolation | 0.005 | 0.74 | 28 | 0.467 |
| 1hr post dose:150 mg | 0.010 | 0.22 | 62 | 0.825 |
| 2.5hr post dose:150 mg | -0.009 | -0.21 | 62 | 0.836 |
| 1hr post dose:300 mg | -0.032 | -0.74 | 62 | 0.465 |
| 2.5hr post dose:300 mg | 0.001 | 0.01 | 62 | 0.989 |
| | **F-tests** | | | |
| | **Numerator DF** | **Denominator DF** | **F-value** | **p-value** |
| Intercept (placebo, baseline) | 1 | 62 | 15889.41 | <.001 |
| Time | 2 | 62 | 3.42 | 0.039 |
| Group | 2 | 28 | 0.50 | 0.610 |
| TICS chronic stress | 1 | 28 | 4.26 | 0.048 |
| Sweets intake | 1 | 28 | 2.32 | 0.139 |
| TICS social overload | 1 | 28 | 0.29 | 0.594 |
| TICS pressure to succeed | 1 | 28 | 5.95 | 0.021 |
| TICS social isolation | 1 | 28 | 0.54 | 0.470 |
| Time:group | 4 | 62 | 0.45 | 0.776 |
| | **Post-hoc Tukey tests for time** | | | |
| | **Estimate** | **z-value** | | **p-value** |
| 1hr post dose - baseline | -0.030 | -1.684 | | 0.211 |
| 2.5hr post dose – baseline | -0.048 | -2.690 | | 0.020 |
| 2.5hr post dose - 1hr post dose | -0.018 | -1.049 | | 0.546 |

**Table 13: Inferential statistics for Spatial Working Memory Accuracy in the ITT population**

| | Linear mixed effects model summary | | | |
|---|---|---|---|---|
| | **Estimate** | **t-value** | **DF** | **p-value** |
| Intercept (placebo, baseline) | 97.035 | 92.81 | 58 | <.001 |
| 1hr post dose | -0.521 | -0.48 | 58 | 0.634 |
| 2.5hr post dose | -1.062 | -0.95 | 58 | 0.346 |
| 150 mg group | -0.001 | 0.00 | 29 | 0.999 |
| 300 mg group | -0.044 | -0.03 | 29 | 0.973 |
| Sleep disruption | -1.756 | -2.27 | 29 | 0.031 |
| TICS lack of acceptation | 0.094 | 0.58 | 29 | 0.564 |
| TICS social tension | 0.16 | 1.10 | 29 | 0.280 |
| 1hr post dose:150 mg | -2.204 | -1.33 | 58 | 0.187 |
| 2.5hr post dose:150 mg | 0.356 | 0.21 | 58 | 0.835 |
| 1hr post dose:300 mg | -1.198 | -0.78 | 58 | 0.439 |
| 2.5hr post dose:300 mg | 0.333 | 0.21 | 58 | 0.832 |
| | **F-tests** | | | |
| | **Numerator DF** | **Denominator DF** | **F-value** | **p-value** |
| Intercept (placebo, baseline) | 1 | 58 | 74529.15 | <.001 |
| Time | 2 | 58 | 2.85 | 0.066 |
| Group | 2 | 29 | 0.29 | 0.749 |
| Sleep disruption | 1 | 29 | 5.53 | 0.026 |
| TICS lack of acceptation | 1 | 29 | 3.46 | 0.073 |
| TICS social tension | 1 | 29 | 1.28 | 0.267 |
| Time:group | 4 | 58 | 0.73 | 0.578 |

**Table 14: Inferential statistics for Spatial Working Memory Reaction Time in the ITT population, outcome was log transformed, one outlier was manually removed**

| | Linear mixed effects model summary | | | |
|---|---|---|---|---|
| | **Estimate** | **t-value** | **DF** | **p-value** |
| Intercept (placebo, baseline) | 6.502 | 122.34 | 56 | <.001 |
| 1hr post dose | -0.069 | -1.90 | 56 | 0.063 |
| 2.5hr post dose | -0.004 | -0.12 | 56 | 0.903 |
| 150 mg group | -0.030 | -0.50 | 31 | 0.618 |
| 300 mg group | -0.023 | -0.39 | 31 | 0.697 |
| Sweets intake | -0.022 | -2.32 | 31 | 0.027 |
| 1hr post dose:150 mg | 0.037 | 0.63 | 56 | 0.529 |
| 2.5hr post dose:150 mg | -0.002 | -0.04 | 56 | 0.970 |
| 1hr post dose:300 mg | 0.065 | 1.22 | 56 | 0.228 |
| 2.5hr post dose:300 mg | -0.050 | -0.95 | 56 | 0.345 |
| | **F-tests** | | | |
| | **Numerator DF** | **Denominator DF** | **F-value** | **p-value** |
| Intercept (placebo, baseline) | 1 | 56 | 98081.33 | <.001 |
| Time | 2 | 56 | 1.30 | 0.281 |
| Group | 2 | 31 | 0.07 | 0.935 |
| Sweets intake | 1 | 31 | 5.39 | 0.027 |
| Time:group | 4 | 56 | 1.26 | 0.297 |

**Table 15: Inferential statistics for Simple Reaction Time in the ITT population**

| | Linear mixed effects model summary | | | |
|---|---|---|---|---|
| | **Estimate** | **t-value** | **DF** | **p-value** |
| Intercept (placebo, baseline) | 423.899 | 11.54 | 63 | <.001 |
| 1hr post dose | 2.037 | 0.33 | 63 | 0.743 |
| 2.5hr post dose | -3.927 | -0.62 | 63 | 0.540 |
| 150 mg group | -10.031 | -0.87 | 29 | 0.394 |
| 300 mg group | -17.293 | -1.51 | 29 | 0.143 |
| Sweets intake | -4.581 | -2.32 | 29 | 0.027 |
| Sleep disruptions | -12.448 | -1.31 | 29 | 0.201 |
| Sleep duration | -6.865 | -1.46 | 29 | 0.155 |
| 1hr post dose:150 mg | 6.765 | 0.76 | 63 | 0.452 |
| 2.5hr post dose:150 mg | 13.212 | 1.46 | 63 | 0.150 |
| 1hr post dose:300 mg | 5.783 | 0.66 | 63 | 0.510 |
| 2.5hr post dose:300 mg | 6.246 | 0.70 | 63 | 0.484 |
| | **F-tests** | | | |
| | **Numerator DF** | **Denominator DF** | **F-value** | **p-value** |
| Intercept (placebo, baseline) | 1 | 63 | 6646.291 | <.001 |
| Time | 2 | 63 | 1.468 | 0.238 |
| Group | 2 | 29 | 1.266 | 0.297 |
| Sweets intake | 1 | 29 | 5.363 | 0.028 |
| Sleep disruptions | 1 | 29 | 3.342 | 0.078 |
| Sleep duration | 1 | 29 | 2.130 | 0.155 |
| Time:group | 4 | 63 | 0.573 | 0.683 |

**Table 16: Inferential statistics for Digit Vigilance Accuracy in the ITT population**

| | Linear mixed effects model summary | | | |
|---|---|---|---|---|
| | Estimate | t-value | DF | p-value |
| Intercept (placebo, baseline) | 92.996 | 49.36 | 60 | <.001 |
| 1hr post dose | -0.606 | -1.05 | 60 | 0.299 |
| 2.5hr post dose | -0.986 | -1.73 | 60 | 0.089 |
| 150 mg group | 0.069 | 0.1 | 30 | 0.921 |
| 300 mg group | 0.759 | 1.11 | 30 | 0.276 |
| Sweets intake | 0.283 | 2.91 | 30 | 0.007 |
| Sleep duration | 0.596 | 2.63 | 30 | 0.013 |
| 1hr post dose:150 mg | -0.605 | -0.74 | 60 | 0.463 |
| 2.5hr post dose:150 mg | -0.203 | -0.25 | 60 | 0.807 |
| 1hr post dose:300 mg | 0.606 | 0.76 | 60 | 0.452 |
| 2.5hr post dose:300 mg | 0.185 | 0.23 | 60 | 0.819 |
| | F-tests | | | |
| | Numerator DF | Denominator DF | F-value | p-value |
| Intercept (placebo, baseline) | 1 | 60 | 231103.2 | <.001 |
| Time | 2 | 60 | 4.47 | 0.016 |
| Group | 2 | 30 | 3.57 | 0.041 |
| Sweets intake | 1 | 30 | 7.50 | 0.010 |
| Sleep duration | 1 | 30 | 6.93 | 0.013 |
| Time:group | 4 | 60 | 0.60 | 0.667 |
| | Post-hoc Tukey tests for time | | | |
| | Estimate | z-value | | p-value |
| 1hr post dose - baseline | -0.5879 | -1.794 | | 0.172 |
| 2.5hr post dose – baseline | -0.9773 | -2.935 | | 0.009 |
| 2.5hr post dose - 1hr post dose | -0.3893 | -1.169 | | 0.472 |
| | Post-hoc Tukey tests for group | | | |
| | Estimate | z-value | | p-value |
| 150 mg group - placebo | -0.08511 | -0.144 | | 0.989 |
| 300 mg group - placebo | 1.12057 | 1.938 | | 0.128 |
| 300 mg group - 150 mg group | 1.20569 | 2.051 | | 0.100 |

**Table 17: Inferential statistics for Digit Vigilance Reaction Time in the ITT population**

| | Linear mixed effects model summary | | | |
|---|---|---|---|---|
| | **Estimate** | **t-value** | **DF** | **p-value** |
| Intercept (placebo, baseline) | 623.566 | 10.83 | 66 | 0 |
| 1hr post dose | -2.22 | -0.28 | 66 | 0.778 |
| 2.5hr post dose | 1.374 | 0.18 | 66 | 0.861 |
| 150 mg group | 8.87 | 0.51 | 31 | 0.613 |
| 300 mg group | 0.995 | 0.06 | 31 | 0.955 |
| Sleep duration | -20.279 | -2.83 | 31 | 0.008 |
| Sweets intake | -6.415 | -2.05 | 31 | 0.049 |
| 1hr post dose:150 mg | 10.893 | 0.98 | 66 | 0.329 |
| 2.5hr post dose:150 mg | 12.497 | 1.13 | 66 | 0.264 |
| 1hr post dose:300 mg | 9.838 | 0.89 | 66 | 0.378 |
| 2.5hr post dose:300 mg | -9.786 | -0.88 | 66 | 0.38 |
| **F-tests** | | | | |
| | **Numerator DF** | **Denominator DF** | **F-value** | **p-value** |
| Intercept (placebo, baseline) | 1 | 66 | 4634.454 | <.001 |
| Time | 2 | 66 | 0.537 | 0.587 |
| Group | 2 | 31 | 0.711 | 0.499 |
| Sleep duration | 1 | 31 | 7.783 | 0.009 |
| Sweets intake | 1 | 31 | 4.189 | 0.049 |
| Time:group | 4 | 66 | 1.631 | 0.177 |

**Table 18: Inferential statistics for Digit Vigilance False Alarms in the ITT population, outcome was log transformed after adding 1 to each value**

| | Linear mixed effects model summary | | | |
|---|---|---|---|---|
| | **Estimate** | **t-value** | **DF** | **p-value** |
| Intercept (placebo, baseline) | 0.407 | 1.30 | 62 | 0.200 |
| 1hr post dose | -0.116 | -0.62 | 62 | 0.536 |
| 2.5hr post dose | -0.058 | -0.31 | 62 | 0.760 |
| 150 mg group | -0.243 | -1.12 | 31 | 0.269 |
| 300 mg group | 0.147 | 0.69 | 31 | 0.495 |
| BFP | 0.017 | 2.35 | 31 | 0.025 |
| Sweets intake | -0.072 | -2.41 | 31 | 0.022 |
| 1hr post dose:150 mg | 0.278 | 1.04 | 62 | 0.303 |
| 2.5hr post dose:150 mg | 0.421 | 1.54 | 62 | 0.129 |
| 1hr post dose:300 mg | -0.293 | -1.12 | 62 | 0.268 |
| 2.5hr post dose:300 mg | -0.274 | -1.03 | 62 | 0.307 |
| | **F-tests** | | | |
| | **Numerator DF** | **Denominator DF** | **F-value** | **p-value** |
| Intercept (placebo, baseline) | 1 | 62 | 106.254 | <.001 |
| Time | 2 | 62 | 0.833 | 0.440 |
| Group | 2 | 31 | 0.084 | 0.920 |
| BFP | 1 | 31 | 9.364 | 0.005 |
| Sweets intake | 1 | 31 | 5.978 | 0.020 |
| Time:group | 4 | 62 | 1.927 | 0.117 |

**Table 19: Inferential statistics for Choice Reaction Time Accuracy in the ITT population**

| | Linear mixed effects model summary | | | |
|---|---|---|---|---|
| | **Estimate** | **t-value** | **DF** | **p-value** |
| Intercept (placebo, baseline) | 97.333 | 105.11 | 66 | <.001 |
| 1hr post dose | -1.500 | -2.03 | 66 | 0.046 |
| 2.5hr post dose | -1.167 | -1.58 | 66 | 0.119 |
| 150 mg group | -2.000 | -1.53 | 33 | 0.136 |
| 300 mg group | -1.333 | -1.02 | 33 | 0.316 |
| 1hr post dose:150 mg | 1.167 | 1.12 | 66 | 0.268 |
| 2.5hr post dose:150 mg | 0.667 | 0.64 | 66 | 0.526 |
| 1hr post dose:300 mg | 0.833 | 0.80 | 66 | 0.428 |
| 2.5hr post dose:300 mg | 0.333 | 0.32 | 66 | 0.751 |
| | **F-tests** | | | |
| | **Numerator DF** | **Denominator DF** | **F-value** | **p-value** |
| Intercept (placebo, baseline) | 1 | 66 | 40648.45 | <.001 |
| Time | 2 | 66 | 2.54 | 0.086 |
| Group | 2 | 33 | 0.74 | 0.483 |
| Time:group | 4 | 66 | 0.34 | 0.851 |

**Table 20: Inferential statistics for Choice Reaction Time in the ITT population**

| | Linear mixed effects model summary | | | |
|---|---|---|---|---|
| | **Estimate** | **t-value** | **DF** | **p-value** |
| Intercept (placebo, baseline) | 371.24 | 14.85 | 63 | <.001 |
| 1hr post dose | -4.593 | -0.77 | 63 | 0.444 |
| 2.5hr post dose | -11.768 | -1.97 | 63 | 0.053 |
| 150 mg group | 3.958 | 0.27 | 28 | 0.792 |
| 300 mg group | 2.906 | 0.18 | 28 | 0.859 |
| BFP | 1.003 | 1.31 | 28 | 0.202 |
| Gender | 12.094 | 0.84 | 28 | 0.407 |
| TICS social overload | 1.848 | 1.05 | 28 | 0.302 |
| TICS chronic worrying | 1.044 | 0.53 | 28 | 0.601 |
| 1hr post dose:150 mg | 11.864 | 1.41 | 63 | 0.162 |
| 2.5hr post dose:150 mg | 14.188 | 1.72 | 63 | 0.091 |
| 1hr post dose:300 mg | -0.156 | -0.02 | 63 | 0.985 |
| 2.5hr post dose:300 mg | -0.133 | -0.02 | 63 | 0.987 |
| | **F-tests** | | | |
| | **Numerator DF** | **Denominator DF** | **F-value** | **p-value** |
| Intercept (placebo, baseline) | 1 | 63 | 6179.861 | <.001 |
| Time | 2 | 63 | 2.575 | 0.084 |
| Group | 2 | 28 | 0.970 | 0.391 |
| BFP | 1 | 28 | 6.748 | 0.015 |
| Gender | 1 | 28 | 0.947 | 0.339 |
| TICS social overload | 1 | 28 | 2.543 | 0.122 |
| TICS chronic worrying | 1 | 28 | 0.270 | 0.607 |
| Time:group | 4 | 63 | 1.171 | 0.332 |

#### 4.3 Physiological outcomes

**Table 21: Inferential statistics for HR in the ITT population, outcome was log transformed**

| | Linear mixed effects model summary | | | |
|---|---|---|---|---|
| | **Estimate** | **t-value** | **DF** | **p-value** |
| Intercept (placebo, baseline) | 4.007 | 43.17 | 177 | <.001 |
| Time | 0.000 | -0.62 | 177 | 0.535 |
| 150 mg group | -0.080 | -1.37 | 32 | 0.181 |
| 300 mg group | 0.007 | 0.13 | 32 | 0.900 |
| Gender | 0.129 | 2.59 | 32 | 0.014 |
| Time:150 mg | 0.001 | 2.65 | 177 | 0.009 |
| Time:300 mg | 0.000 | 0.87 | 177 | 0.386 |
| | **F-tests** | | | |
| | **Numerator DF** | **Denominator DF** | **F-value** | **p-value** |
| Intercept (placebo, baseline) | 1 | 177 | 32709.81 | <.001 |
| Time | 1 | 177 | 3.23 | 0.074 |
| Group | 2 | 32 | 0.32 | 0.727 |
| Gender | 1 | 32 | 6.72 | 0.014 |
| Time:group | 2 | 177 | 3.65 | 0.028 |

**Table 22: Inferential statistics for diastolic BP in the ITT population**

| Linear mixed effects model summary | | | | |
|---|---|---|---|---|
| | **Estimate** | **t-value** | **DF** | **p-value** |
| Intercept (placebo, baseline) | 58.222 | 10.71 | 177 | <.001 |
| Time | -0.006 | -0.86 | 177 | 0.392 |
| 150 mg group | 1.173 | 0.42 | 31 | 0.678 |
| 300 mg group | 0.587 | 0.21 | 31 | 0.834 |
| BFP | 0.22 | 1.59 | 31 | 0.122 |
| Weight | 0.114 | 1.56 | 31 | 0.13 |
| Time:150 mg | 0.009 | 0.86 | 177 | 0.393 |
| Time:300 mg | 0.008 | 0.77 | 177 | 0.442 |
| **F-tests** | | | | |
| | **Numerator DF** | **Denominator DF** | **F-value** | **p-value** |
| Intercept (placebo, baseline) | 1 | 177 | 4552.855 | <.001 |
| Time | 1 | 177 | 0.025 | 0.876 |
| Group | 2 | 31 | 0.311 | 0.735 |
| BFP | 1 | 31 | 6.736 | 0.014 |
| Weight | 1 | 31 | 2.424 | 0.130 |
| Time:group | 2 | 177 | 0.445 | 0.642 |

**Table 23: Inferential statistics for systolic BP in the ITT population**

| | Linear mixed effects model summary | | | |
|---|---|---|---|---|
| | **Estimate** | **t-value** | **DF** | **p-value** |
| Intercept (placebo, baseline) | 101.74 | 3.21 | 176 | 0.002 |
| Time | 0.013 | 1.38 | 176 | 0.17 |
| 150 mg group | -2.411 | -0.62 | 30 | 0.539 |
| 300 mg group | 0.044 | 0.01 | 30 | 0.991 |
| Gender | -6.938 | -1.24 | 30 | 0.224 |
| WHR | 37.601 | 1.19 | 30 | 0.243 |
| Fruit and vegetable intake | -1.04 | -1.03 | 30 | 0.313 |
| Time:150 mg | 0.02 | 1.53 | 176 | 0.128 |
| Time:300 mg | 0.01 | 0.60 | 176 | 0.551 |
| | **F-tests** | | | |
| | **Numerator DF** | **Denominator DF** | **F-value** | **p-value** |
| Intercept (placebo, baseline) | 1 | 176 | 5743.48 | <.001 |
| Time | 1 | 176 | 16.83 | <.001 |
| Group | 2 | 30 | 0.09 | 0.913 |
| Gender | 1 | 30 | 18.14 | <.001 |
| WHR | 1 | 30 | 1.22 | 0.278 |
| Fruit and vegetable intake | 1 | 30 | 0.93 | 0.344 |
| Time:group | 2 | 176 | 1.19 | 0.308 |
| | **Linear mixed effects model summary without interaction** | | | |
| | **Estimate** | **t-value** | **DF** | **p-value** |
| Intercept (placebo, baseline) | 101.165 | 3.19 | 178 | 0.002 |
| Time | 0.022 | 4.10 | 178 | <.001 |
| 150 mg group | -1.040 | -0.28 | 30 | 0.785 |
| 300 mg group | 0.571 | 0.15 | 30 | 0.880 |
| Gender | -6.959 | -1.24 | 30 | 0.223 |
| WHR | 37.575 | 1.19 | 30 | 0.244 |
| Fruit and vegetable intake | -1.038 | -1.02 | 30 | 0.314 |

**Table 24: Inferential statistics for salivary cortisol in the ITT population, outcome was log transformed**

| | Linear mixed effects model summary | | | |
|---|---|---|---|---|
| | **Estimate** | **t-value** | **DF** | **p-value** |
| Intercept (placebo, baseline) | 1.509 | 3.54 | 171 | <.001 |
| Time | -0.001 | -2.54 | 171 | 0.012 |
| 150 mg group | -0.268 | -1.97 | 29 | 0.059 |
| 300 mg group | -0.249 | -1.79 | 29 | 0.085 |
| Weight | -0.003 | -0.79 | 29 | 0.433 |
| Sleep quality | -0.049 | -1.27 | 29 | 0.215 |
| Sweets intake | 0.026 | 0.95 | 29 | 0.348 |
| Level of activity | 0.073 | 2.62 | 29 | 0.014 |
| Time:150 mg | 0.001 | 0.66 | 171 | 0.512 |
| Time:300 mg | 0.001 | 1.51 | 171 | 0.133 |
| | **F-tests** | | | |
| | **Numerator DF** | **Denominator DF** | **F-value** | **p-value** |
| Intercept (placebo, baseline) | 1 | 171 | 478.73 | <.001 |
| Time | 1 | 171 | 6.81 | 0.010 |
| Group | 2 | 29 | 1.82 | 0.180 |
| weight | 1 | 29 | 3.71 | 0.064 |
| Sleep quality | 1 | 29 | 2.06 | 0.162 |
| Sweets intake | 1 | 29 | 2.00 | 0.168 |
| Level of activity | 1 | 29 | 6.38 | 0.017 |
| Time:group | 2 | 171 | 1.14 | 0.321 |

**Table 25: Inferential statistics for salivary oxytocin in the ITT population**

| | Linear mixed effects model summary | | | |
|---|---|---|---|---|
| | **Estimate** | **t-value** | **DF** | **p-value** |
| Intercept (placebo, before treatment) | 1.429 | 9.99 | 31 | <.001 |
| Time (after treatment) | -0.105 | -1.11 | 31 | 0.278 |
| 150 mg group | 0.496 | 2.96 | 31 | 0.006 |
| 300 mg group | 0.189 | 1.11 | 31 | 0.275 |
| CAR increase | -0.010 | -1.11 | 31 | 0.274 |
| Time:150 mg | -0.097 | -0.71 | 31 | 0.483 |
| Time:300 mg | 0.200 | 1.45 | 31 | 0.156 |
| | **F-tests** | | | |
| | **Numerator DF** | **Denominator DF** | **F-value** | **p-value** |
| Intercept (placebo, baseline) | 1 | 31 | 643.08 | <.001 |
| Time | 1 | 31 | 1.52 | 0.226 |
| Group | 2 | 31 | 5.59 | 0.009 |
| CAR increase | 1 | 31 | 1.17 | 0.287 |
| Time:group | 2 | 31 | 2.34 | 0.113 |
| | **Post-hoc Tukey tests for group** | | | |
| | **Estimate** | **z-value** | | **p-value** |
| 150 mg group – placebo group | 0.444 | 2.920 | | 0.010 |
| 300 mg group – placebo group | 0.290 | 1.858 | | 0.151 |
| 300 mg group – 150 mg group | -0.154 | -1.016 | | 0.567 |

##### 4.4 Exploratory cognitive outcomes

**Table 26: Inferential statistics for cognitive factor: secondary memory in the ITT population**

| | Linear mixed effects model summary | | | |
|---|---|---|---|---|
| | **Estimate** | **t-value** | **DF** | **p-value** |
| Intercept (placebo, baseline) | 324.695 | 12.51 | 66 | <.001 |
| 1hr post dose | -33.808 | -3.58 | 66 | 0.001 |
| 2.5hr post dose | -26.38 | -2.79 | 66 | 0.007 |
| 150 mg group | -20.387 | -1.31 | 32 | 0.199 |
| 300 mg group | -9.481 | -0.61 | 32 | 0.546 |
| Weight | -0.598 | -1.83 | 32 | 0.076 |
| 1hr post dose:150 mg | 22.538 | 1.69 | 66 | 0.096 |
| 2.5hr post dose:150 mg | 0.627 | 0.05 | 66 | 0.963 |
| 1hr post dose:300 mg | 17.419 | 1.3 | 66 | 0.197 |
| 2.5hr post dose:300 mg | 19.99 | 1.5 | 66 | 0.139 |
| | **F-tests** | | | |
| | **Numerator DF** | **Denominator DF** | **F-value** | **p-value** |
| Intercept (placebo, baseline) | 1 | 66 | 2211.45 | <.001 |
| Time | 2 | 66 | 8.97 | <.001 |
| Group | 2 | 32 | 0.66 | 0.523 |
| Weight | 1 | 32 | 3.36 | 0.076 |
| Time:group | 4 | 66 | 1.68 | 0.165 |
| | **Post-hoc Tukey tests for time** | | | |
| | **Estimate** | **z-value** | | **p-value** |
| 1hr post dose - baseline | 20.49 | -3.685 | | <.001 |
| 2.5hr post dose – baseline | 19.51 | -3.508 | | 0.001 |
| 2.5hr post dose - 1hr post dose | 0.98 | 0.176 | | 0.983 |

**Table 27: Inferential statistics for cognitive factor: Working Memory in the ITT population**

| | Linear mixed effects model summary | | | |
|---|---|---|---|---|
| | **Estimate** | **t-value** | **DF** | **p-value** |
| Intercept (placebo, baseline) | 197.619 | 78.36 | 59 | <.001 |
| 1hr post dose | -0.243 | -0.14 | 59 | 0.889 |
| 2.5hr post dose | -1.554 | -0.87 | 59 | 0.390 |
| 150 mg group | -2.910 | -1.14 | 31 | 0.263 |
| 300 mg group | 2.215 | 0.90 | 31 | 0.376 |
| Sleep disruption | -3.579 | -1.90 | 31 | 0.067 |
| TICS work dissatisfaction | -0.346 | -1.89 | 31 | 0.069 |
| 1hr post dose:150 mg | -0.530 | -0.20 | 59 | 0.841 |
| 2.5hr post dose:150 mg | 2.271 | 0.85 | 59 | 0.398 |
| 1hr post dose:300 mg | 0.191 | 0.08 | 59 | 0.939 |
| 2.5hr post dose:300 mg | 1.381 | 0.55 | 59 | 0.583 |
| | **F-tests** | | | |
| | **Numerator DF** | **Denominator DF** | **F-value** | **p-value** |
| Intercept (placebo, baseline) | 1 | 59 | 54527.41 | <.001 |
| Time | 2 | 59 | 0.06 | 0.939 |
| Group | 2 | 31 | 2.34 | 0.113 |
| Sleep disruption | 1 | 31 | 6.29 | 0.018 |
| TICS work dissatisfaction | 1 | 31 | 3.47 | 0.072 |
| Time:group | 4 | 59 | 0.32 | 0.864 |

**Table 28: Inferential statistics for cognitive factor: Speed of Memory in the ITT population**

| | Linear mixed effects model summary | | | |
|---|---|---|---|---|
| | **Estimate** | **t-value** | **DF** | **p-value** |
| Intercept (placebo, baseline) | 3115.71 | 23.5 | 66 | <.000 |
| 1hr post dose | -111.264 | -1.54 | 66 | 0.129 |
| 2.5hr post dose | -100.341 | -1.39 | 66 | 0.17 |
| 150 mg group | 295.174 | 2.03 | 31 | 0.051 |
| 300 mg group | 8.126 | 0.05 | 31 | 0.957 |
| Sleep disruption | -324.892 | -2.72 | 31 | 0.011 |
| TICS: social isolation | 42.317 | 3.48 | 31 | 0.002 |
| 1hr post dose:150 mg | -57.637 | -0.56 | 66 | 0.575 |
| 2.5hr post dose:150 mg | -159.307 | -1.56 | 66 | 0.124 |
| 1hr post dose:300 mg | 58.047 | 0.57 | 66 | 0.572 |
| 2.5hr post dose:300 mg | 14.038 | 0.14 | 66 | 0.891 |
| | | **F-tests** | | |
| | **Numerator DF** | **Denominator DF** | **F-value** | **p-value** |
| Intercept (placebo, baseline) | 1 | 66 | 3446.38 | <.000 |
| Time | 2 | 66 | 6.86 | 0.002 |
| Group | 2 | 31 | 0.69 | 0.510 |
| Sleep disruption | 1 | 31 | 5.01 | 0.033 |
| TICS: social isolation | 1 | 31 | 12.12 | 0.002 |
| Time:group | 4 | 66 | 0.97 | 0.431 |
| | **Post-hoc Tukey tests for time** | | | |
| | **Estimate** | **z-value** | | **p-value** |
| 1hr post dose - baseline | -111.13 | -2.663 | | 0.021 |
| 2.5hr post dose – baseline | -148.76 | -3.564 | | 0.001 |
| 2.5hr post dose - 1hr post dose | -37.64 | -0.902 | | 0.639 |

**Table 29: Inferential statistics for cognitive factor: Accuracy of Attention in the ITT population**

| | Linear mixed effects model summary | | | |
|---|---|---|---|---|
| | **Estimate** | **t-value** | **DF** | **p-value** |
| Intercept (placebo, baseline) | 96.556 | 130.12 | 59 | <.001 |
| 1hr post dose | -0.845 | -1.65 | 59 | 0.105 |
| 2.5hr post dose | -0.768 | -1.54 | 59 | 0.128 |
| 150 mg group | 0.122 | 0.15 | 31 | 0.886 |
| 300 mg group | 0.266 | 0.33 | 31 | 0.745 |
| Sweets intake | 0.305 | 2.26 | 31 | 0.031 |
| 1hr post dose:150 mg | -0.516 | -0.68 | 59 | 0.499 |
| 2.5hr post dose:150 mg | -1.080 | -1.44 | 59 | 0.154 |
| 1hr post dose:300 mg | 0.473 | 0.65 | 59 | 0.518 |
| 2.5hr post dose:300 mg | 0.055 | 0.08 | 59 | 0.940 |
| | **F-tests** | | | |
| | **Numerator DF** | **Denominator DF** | **F-value** | **p-value** |
| Intercept (placebo, baseline) | 1 | 59 | 115790.5 | <.001 |
| Time | 2 | 59 | 6.75 | 0.002 |
| Group | 2 | 31 | 0.54 | 0.588 |
| Sweets intake | 1 | 31 | 5.15 | 0.030 |
| Time:group | 4 | 59 | 0.83 | 0.509 |
| | **Post-hoc Tukey tests for time** | | | |
| | **Estimate** | **z-value** | | **p-value** |
| 1hr post dose - baseline | -0.816 | -2.695 | | 0.019 |
| 2.5hr post dose – baseline | -1.067 | -3.528 | | 0.001 |
| 2.5hr post dose - 1hr post dose | -0.252 | -0.850 | | 0.672 |

**Table 30: Inferential statistics for cognitive factor: Speed of Attention in the ITT population**

| | Linear mixed effects model summary | | | |
|---|---|---|---|---|
| | **Estimate** | **t-value** | **DF** | **p-value** |
| Intercept (placebo, baseline) | 1543.576 | 12.51 | 64 | <.001 |
| 1hr post dose | -6.297 | -0.51 | 64 | 0.613 |
| 2.5hr post dose | -3.099 | -0.25 | 64 | 0.803 |
| 150 mg group | -8.215 | -0.23 | 29 | 0.823 |
| 300 mg group | -7.004 | -0.19 | 29 | 0.848 |
| Sweets intake | -13.589 | -2.05 | 29 | 0.050 |
| Sleep disruption | -58.679 | -1.83 | 29 | 0.077 |
| Sleep duration | -28.798 | -1.82 | 29 | 0.079 |
| 1hr post dose:150 mg | 29.674 | 1.66 | 64 | 0.103 |
| 2.5hr post dose:150 mg | 31.48 | 1.76 | 64 | 0.084 |
| 1hr post dose:300 mg | 16.986 | 0.97 | 64 | 0.336 |
| 2.5hr post dose:300 mg | -14.895 | -0.85 | 64 | 0.399 |
| | **F-tests** | | | |
| | **Numerator DF** | **Denominator DF** | **F-value** | **p-value** |
| Intercept (placebo, baseline) | 1 | 64 | 7688.12 | <.001 |
| Time | 2 | 64 | 0.84 | 0.437 |
| Group | 2 | 29 | 0.23 | 0.798 |
| Sweets intake | 1 | 29 | 4.16 | 0.051 |
| Sleep disruption | 1 | 29 | 6.13 | 0.019 |
| Sleep duration | 1 | 29 | 3.30 | 0.080 |
| Time:group | 4 | 64 | 2.38 | 0.061 |

**Table 31: Inferential statistics for Immediate Word Recall Accuracy for combined treatment group in the ITT population**

| | Linear mixed effects model summary | | | |
|---|---|---|---|---|
| | Estimate | t-value | DF | p-value |
| Intercept (placebo, baseline) | 71.446 | 5.44 | 68 | <.001 |
| 1hr post dose | -15.357 | -4.17 | 68 | <.001 |
| 2.5hr post dose | -6.189 | -1.68 | 68 | 0.097 |
| sage group(collapsed) | -5.303 | -1.14 | 31 | 0.264 |
| Weight | -0.171 | -1.5 | 31 | 0.145 |
| Gender | 5.571 | 1.35 | 31 | 0.188 |
| TICS social overload | -0.911 | -2.03 | 31 | 0.051 |
| 1hr post dose:sage group(collapsed) | 12.658 | 2.81 | 68 | 0.007 |
| 2.5hr post dose:sage group(collapsed) | 3.887 | 0.86 | 68 | 0.392 |
| | F-tests | | | |
| | Numerator DF | Denominator DF | F-value | p-value |
| Intercept (placebo, baseline) | 1 | 68 | 958.98 | <.001 |
| Time | 2 | 68 | 5.30 | 0.007 |
| group | 1 | 31 | 0.14 | 0.714 |
| Weight | 1 | 31 | 4.83 | 0.036 |
| Gender | 1 | 31 | 1.04 | 0.315 |
| TICS social overload | 1 | 31 | 4.14 | 0.051 |
| Time:group | 2 | 68 | 4.14 | 0.020 |

**Table 32: Inferential statistics for HR for combined treatment group in the ITT population, outcome was log transformed**

| | Linear mixed effects model summary | | | |
|---|---|---|---|---|
| | Estimate | t-value | DF | p-value |
| Intercept (placebo, baseline) | 4.021 | 43.51 | 178 | <.001 |
| Time | 0.000 | -0.62 | 178 | 0.538 |
| sage group(collapsed) | -0.036 | -0.71 | 33 | 0.480 |
| Gender | 0.121 | 2.45 | 33 | 0.020 |
| Time:sage group(collapsed) | 0.000 | 2.02 | 178 | 0.045 |
| | F-tests | | | |
| | Numerator DF | Denominator DF | F-value | p-value |
| Intercept (placebo, baseline) | 1 | 178 | 32460.32 | <.001 |
| Time | 1 | 178 | 3.19 | 0.076 |
| Group | 1 | 33 | 0.08 | 0.785 |
| Gender | 1 | 33 | 5.99 | 0.020 |
| Time:group | 1 | 178 | 4.08 | 0.045 |

**Table 33: Inferential statistics for VAS stress for combined treatment group in the ITT population, outcome was square root transformed**

| | Linear mixed effects model summary | | | |
|---|---|---|---|---|
| | **Estimate** | **t-value** | **DF** | **p-value** |
| Intercept (placebo, baseline) | 5.26 | 2.99 | 171 | 0.003 |
| Time | -0.009 | -5.34 | 171 | <.001 |
| sage group(collapsed) | -0.863 | -1.42 | 32 | 0.165 |
| recovery | -0.315 | -1.56 | 32 | 0.129 |
| Sweets intake | 0.164 | 1.21 | 32 | 0.236 |
| Time:sage group(collapsed) | 0.006 | 2.68 | 171 | 0.008 |
| | **F-tests** | | | |
| | **Numerator DF** | **Denominator DF** | **F-value** | **p-value** |
| Intercept (placebo, baseline) | 1 | 171 | 93.12 | <.001 |
| Time | 1 | 171 | 28.90 | <.001 |
| Group | 1 | 32 | 0.45 | 0.507 |
| Gender | 1 | 32 | 4.10 | 0.051 |
| Fruit and vegetable intake | 1 | 32 | 1.46 | 0.235 |
| Time:group | 1 | 171 | 7.18 | 0.008 |

**Table 34: Inferential statistics for SAM dominance for combined treatment group in the ITT population, outcome was square transformed**

| | Linear mixed effects model summary | | | |
|---|---|---|---|---|
| | **Estimate** | **t-value** | **DF** | **p-value** |
| Intercept (placebo, baseline) | 75.097 | 10.47 | 170 | <.001 |
| Time | 0.041 | 3.47 | 170 | 0.001 |
| sage group(collapsed) | -3.957 | -0.72 | 31 | 0.479 |
| Sleep disruptions | -5.033 | -0.87 | 31 | 0.390 |
| TICS work dissatisfaction | -1.507 | -2.68 | 31 | 0.012 |
| Time:sage group(collapsed) | -0.020 | -1.34 | 170 | 0.181 |
| | **F-tests** | | | |
| | **Numerator DF** | **Denominator DF** | **F-value** | **p-value** |
| Intercept (placebo, baseline) | 1 | 170 | 478.54 | <.001 |
| Time | 1 | 170 | 16.61 | <.001 |
| Group | 1 | 31 | 1.46 | 0.236 |
| Sleep disruptions | 1 | 31 | 2.81 | 0.104 |
| TICS work dissatisfaction | 1 | 31 | 7.20 | 0.012 |
| Time:group | 1 | 170 | 1.81 | 0.181 |

**Table 35: Inferential statistics for SAM mood for combined treatment group in the ITT population, outcome was log transformed**

| | Linear mixed effects model summary | | | |
|---|---|---|---|---|
| | **Estimate** | **t-value** | **DF** | **p-value** |
| Intercept (placebo, baseline) | 0.837 | 1.96 | 176 | 0.052 |
| Time | -0.006 | -2.3 | 29 | 0.029 |
| sage group(collapsed) | 0.098 | 1.91 | 29 | 0.066 |
| Weight | 0.205 | 1.98 | 29 | 0.057 |
| Sleep duration | -0.047 | -1.43 | 29 | 0.162 |
| Sleep disruption | 0.023 | 1.91 | 29 | 0.066 |
| recovery | -0.001 | -2.41 | 176 | 0.017 |
| TICS chronic worrying | -0.088 | -0.88 | 29 | 0.386 |
| Time: sage group(collapsed) | 0.001 | 1.44 | 176 | 0.151 |
| | **F-tests** | | | |
| | **Numerator DF** | **Denominator DF** | **F-value** | **p-value** |
| Intercept (placebo, baseline) | 1 | 176 | 587.91 | <.001 |
| Time | 1 | 29 | 4.91 | 0.035 |
| Group | 1 | 29 | 9.24 | 0.005 |
| Weight | 1 | 29 | 5.08 | 0.032 |
| Sleep duration | 1 | 29 | 2.95 | 0.097 |
| Sleep disruption | 1 | 29 | 3.43 | 0.074 |
| recovery | 1 | 176 | 4.59 | 0.034 |
| TICS chronic worrying | 1 | 29 | 0.24 | 0.628 |
| Time:group | 1 | 176 | 2.08 | 0.151 |

**Table 36: Inferential statistics for Picture Recognition Original Stimuli Accuracy in the ITT population, outcome was square transformed**

| | Linear mixed effects model summary | | | |
|---|---|---|---|---|
| | **Estimate** | **t-value** | **DF** | **p-value** |
| Intercept (placebo, baseline) | 6277.332 | 7.47 | 65 | <.001 |
| 1hr post dose | -29.167 | -0.06 | 65 | 0.952 |
| 2.5hr post dose | 664.583 | 1.39 | 65 | 0.170 |
| 150 mg | 62.757 | 0.10 | 30 | 0.918 |
| 300 mg | -96.489 | -0.15 | 30 | 0.879 |
| Gender | 820.047 | 2.02 | 30 | 0.052 |
| TICS social overload | 204.861 | 3.80 | 30 | 0.001 |
| TICS work dissatisfaction | -182.257 | -4.08 | 30 | <.001 |
| 1hr post dose:150 mg | -322.917 | -0.48 | 65 | 0.635 |
| 2.5hr post dose:150 mg | -437.500 | -0.65 | 65 | 0.521 |
| 1hr post dose:300 mg | 511.529 | 0.74 | 65 | 0.460 |
| 2.5hr post dose:300 mg | -105.137 | -0.15 | 65 | 0.879 |

| | F-tests | | | |
|---|---|---|---|---|
| | **Numerator DF** | **Denominator DF** | **F-value** | **p-value** |
| Intercept (placebo, baseline) | 1 | 65 | 1639.5596 | <.001 |
| Time | 2 | 65 | 1.7595 | 0.180 |
| Group | 2 | 30 | 0.2984 | 0.744 |
| Gender | 1 | 30 | 4.9012 | 0.035 |
| TICS social overload | 1 | 30 | 4.9184 | 0.034 |
| TICS work dissatisfaction | 1 | 30 | 16.4233 | <.001 |
| Time:group | 4 | 65 | 0.4817 | 0.749 |

**Table 37: Inferential statistics for Picture Recognition New Stimuli Accuracy in the ITT population**

| | Linear mixed effects model summary | | | |
|---|---|---|---|---|
| | **Estimate** | **t-value** | **DF** | **p-value** |
| Intercept (placebo, baseline) | 72.90881 | 17.49 | 66 | <.001 |
| 1hr post dose | 0.83333 | 0.27 | 66 | 0.787 |
| 2.5hr post dose | 1.66667 | 0.54 | 66 | 0.589 |
| 150 mg | 3.67950 | 0.85 | 31 | 0.400 |
| 300 mg | 3.38297 | 0.76 | 31 | 0.453 |
| CAR increase | 0.37517 | 1.85 | 31 | 0.075 |
| TICS social overload | 0.76488 | 2.16 | 31 | 0.039 |
| 1hr post dose:150 mg | -2.91667 | -0.67 | 66 | 0.504 |
| 2.5hr post dose:150 mg | -9.58333 | -2.21 | 66 | 0.031 |
| 1hr post dose:300 mg | -2.08333 | -0.48 | 66 | 0.633 |
| 2.5hr post dose:300 mg | -1.66667 | -0.38 | 66 | 0.702 |
| | **F-tests** | | | |
| | **Numerator DF** | **Denominator DF** | **F-value** | **p-value** |
| Intercept (placebo, baseline) | 1 | 66 | 3578.161 | <.001 |
| Time | 2 | 66 | 0.700 | 0.500 |
| Group | 2 | 31 | 0.798 | 0.459 |
| Car increase | 1 | 31 | 3.520 | 0.070 |
| TICS social overload | 1 | 31 | 4.665 | 0.039 |
| Time:group | 4 | 66 | 1.565 | 0.194 |
