## Supplementary methods S5 for "Assessment of the Effects of a Sage (*Salvia officinalis*) Extract on Cognitive Performance in Adolescents and Young Adults"

### Inclusion/Exclusion criteria

#### Adolescents

##### *Inclusion criteria*

- healthy male and female individuals
- age group A: 12-14 years
- willing to participate and signed informed consent
- legal representative signed informed consent
- speaks German fluently
- able to understand the background and the purpose of the study, including possible risks and side-effects
- willing to comply with the protocol and the study specific limitations

##### *Exclusion criteria*

- any known current/acute or chronic physical or psychological diseases (e.g. attention deficit (hyperactivity) disorder (AD(H)D), anxiety disorders, diabetes) besides minor medical conditions (e.g. seasonal allergies)
- hypotension (systolic <100; diastolic <60) (except for those whose blood pressure is stable using medication for more than 3 months)
- known hyper- or hypothyroidism unless treated and under control (stable for more than 3 months) intake of any medication during 4 weeks before Visit (V) 1 and during study conduction, which may affect the cognitive performance (e.g. psychotropic, sedating or stimulating medication)
- intake of any medication during 4 weeks before Visit (V) 1 and during study conduction, which may affect the cognitive performance (e.g. psychotropic, sedating or stimulating medication)
- intake of dietary supplements or homoeopathic remedies during 2 weeks before V1 and during the study conduction
- females pregnant or lactating or planning a pregnancy during study conduction
- known dyslexia
- any known vision impairment not corrected by glasses, contact lenses, etc.
- any known addiction to drugs and/or alcohol
- excessive caffeine consumption (>400 mg caffeine/day or V4 cups of caffeinated coffee)
- intake of illegal drugs during 3 weeks before V1 and during study conduction (e.g. cannabis, cocaine)
- any known allergies to the IP or placebo
- smoker
- on a strict diet or practicing sport, extensively
- employee of the sponsor or CRO
- Investigator doubts truthfulness of self-reported health information
- Current participation in another clinical study

#### Young adults

##### *Inclusion criteria*

- healthy male and female individuals
- age group B: 18-25 years
- willing to participate and signed informed consent (legal representative signed informed consent for 12-14 years)
- speaks German fluently
- able to understand the background and the purpose of the study, including possible risks and side-effects
- willing to comply with the protocol and the study specific limitations

##### *Exclusion criteria*

- any known current/acute or chronic physical or psychological diseases (e.g. attention deficit (hyperactivity) disorder (AD(H)D), anxiety disorders, diabetes) besides minor medical conditions (e.g. seasonal allergies)
- hypotension (systolic <100; diastolic <60) (except for those whose blood pressure is stable using medication for more than 3 months)
- known hyper- or hypothyroidism unless treated and under control (stable for more than 3 months) intake of any medication during 4 weeks before Visit (V) 1 and during study conduction, which may affect the cognitive performance (e.g. psychotropic, sedating or stimulating medication)
- intake of dietary supplements or homoeopathic remedies during 2 weeks before V1 and during the study conduction
- females pregnant or lactating or planning a pregnancy during study conduction
- known dyslexia
- any known vision impairment not corrected by glasses, contact lenses, etc.
- any known addiction to drugs and/or alcohol
- excessive caffeine consumption (>400 mg caffeine/day or ≥4 cups of caffeinated coffee)
- intake of illegal drugs during 3 weeks before V1 and during study conduction (e.g. cannabis, cocaine)
- any known allergies to the IP or placebo
- smoker
- on a strict diet or practicing sport, extensively
- employee of the sponsor or CRO
- Investigator doubts truthfulness of self-reported health information
- Current participation in another clinical study

#### Summary of study participants

##### Adolescents

A total of 42 individuals were screened for the study of which one individual met an exclusion criterion and further four individuals were not included due to other reasons. A total of 37 participants entered the study and 36 participants completed the study (see flow chart below).

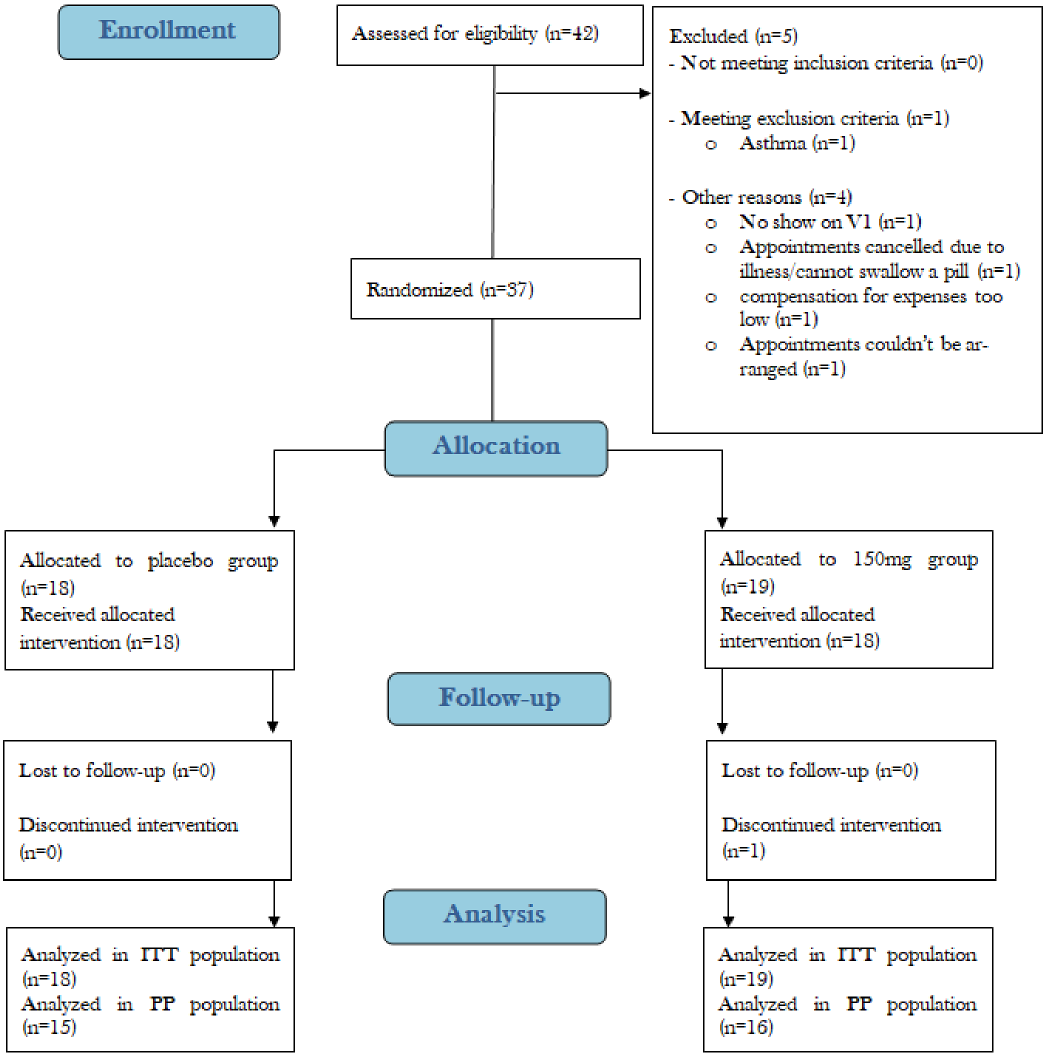

Descriptive statistics for demography and other baseline characteristics in adolescent study population

| | Age [years] | Size [cm] | BMI [kg/m$^2$] | Weight [kg] | WHR | BFP [%] | Pulse rate [bpm] | Systolic blood pressure [mmHg] | Diastolic blood pressure [mmHg] |
|---|---|---|---|---|---|---|---|---|---|
| Mean | 12.95 | 164.89 | 20.21 | 55.17 | 0.8 | 19.46 | 75.86 | 113.65 | 67.89 |
| Median | 13 | 165 | 20.4 | 55.8 | 0.8 | 17.4 | 73 | 111 | 67 |
| SD | 0.78 | 7.67 | 2.99 | 10.21 | 0.05 | 7.68 | 12.38 | 9.82 | 7.31 |
| 95 % CI upper bound | 13.21 | 167.45 | 21.2 | 58.57 | 0.82 | 22.02 | 79.99 | 116.92 | 70.33 |
| 95 % CI lower bound | 12.69 | 162.34 | 19.21 | 51.77 | 0.78 | 16.9 | 71.74 | 110.38 | 65.46 |
| Minimum | 12 | 146 | 15.5 | 37 | 0.7 | 6.8 | 60 | 94 | 56 |
| Maximum | 14 | 184 | 27 | 71.7 | 0.9 | 35.4 | 109 | 134 | 89 |
| N | 37 | 37 | 37 | 37 | 37 | 37 | 37 | 37 | 37 |

Descriptive statistics for gender

| | **Placebo** | **150 mg *Salvia Officinalis L.*** |
|---|---|---|
| Female | 6 | 5 |
| Male | 12 | 14 |
| Percentage female | 33.33% | 26.32% |

#### Young Adults

A total of 51 individuals were screened for the study of which 15 were not included because they did not meet the inclusion criteria, 9 individuals did not enter the study because they did meet an exclusion criteria and further 6 individuals were not included due to other reasons (see flow chart summary below).

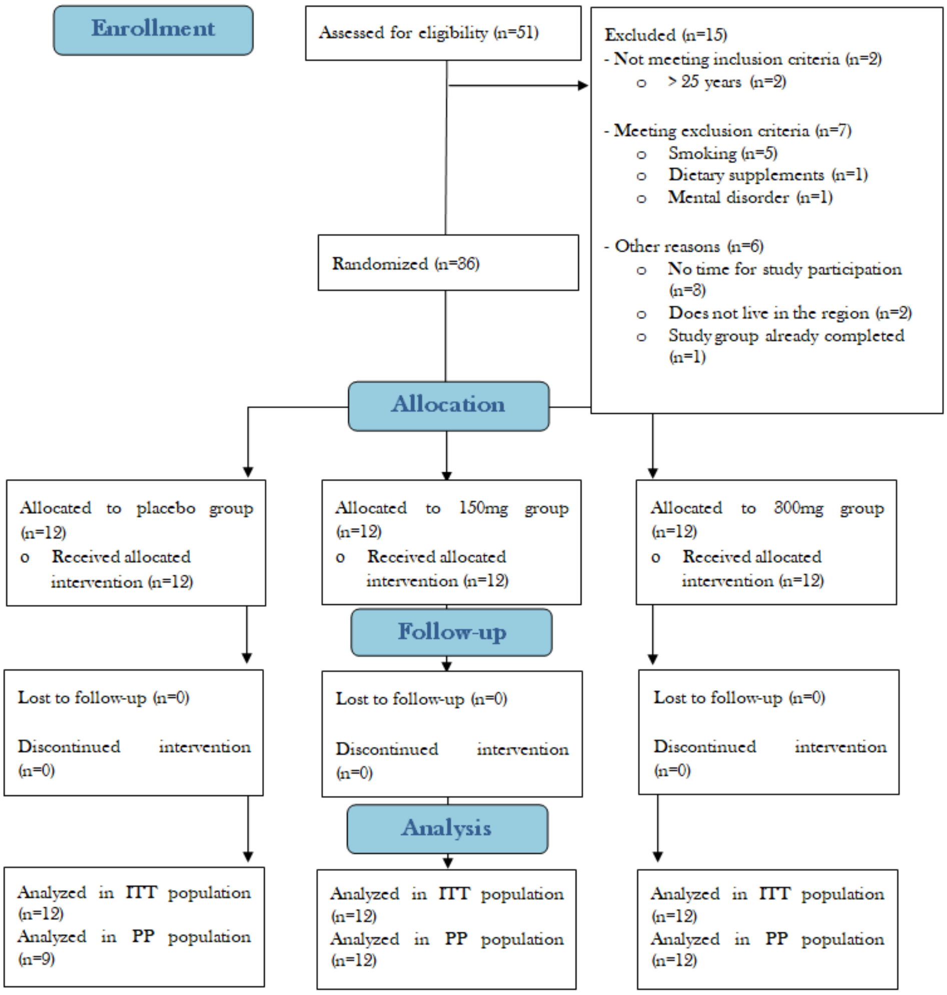

Descriptive statistics for demography and other baseline characteristics in young adult study population

| | Age [years] | Size [cm] | BMI [kg/m$^2$] | Weight [kg] | WHR | BFP [%] | Pulse rate [bpm] | Systolic blood pressure [mmHg] | Diastolic blood pressure [mmHg] |
|---|---|---|---|---|---|---|---|---|---|
| Mean | 22.06 | 169.31 | 24.73 | 70.95 | 0.78 | 31.93 | 75.47 | 122.22 | 77.44 |
| Median | 22 | 167.5 | 23.4 | 66.15 | 0.76 | 33.25 | 77 | 123.5 | 76.5 |
| SD | 1.8 | 9.57 | 5.83 | 17.16 | 0.08 | 9.21 | 12.06 | 12.72 | 8.71 |
| 95 % CI upper bound | 22.67 | 172.54 | 26.7 | 76.75 | 0.81 | 35.04 | 79.55 | 126.52 | 80.39 |
| 95 % CI lower bound | 21.45 | 166.07 | 22.75 | 65.15 | 0.76 | 28.81 | 71.4 | 117.92 | 74.5 |
| Minimum | 18 | 154 | 18.6 | 47.5 | 0.68 | 12.7 | 46 | 97 | 58 |
| Maximum | 25 | 188 | 47.6 | 127.9 | 0.97 | 56.3 | 100 | 148 | 91 |
| N | 36 | 36 | 36 | 36 | 36 | 36 | 36 | 36 | 36 |

Descriptive statistics for gender

| | **Placebo** | **150 mg *Salvia Officinalis L.*** | **300 mg *Salvia Officinalis L.*** |
|---|---|---|---|
| Female | 8 | 9 | 7 |
| Male | 4 | 3 | 5 |
| Percentage female | 66% | 75% | 58% |

### CogTrack™ task descriptions

#### Attention, Concentration, Vigilance

##### *Simple Reaction Time*

This task assesses alertness and the ability to focus concentration by measuring the speed with which a simple motor response can be made to an imminent and expected stimulus, which may occur at any moment. During the task, the word YES is presented in the center of the screen at brief but unpredictable intervals. The participant is instructed to place the right forefinger lightly on the RIGHT arrow keyboard key and to press the key as quickly as possible to the occurrence of the stimulus. It is emphasized that the finger should not be removed from the key between stimuli and that the speed of response is crucially important. It is also made clear that only the word YES will appear and it will always appear in the middle of the screen. Each stimulus remains on the screen until the RIGHT arrow keyboard key is pressed. 50 stimuli are used with randomly varying intervals between 1 and 3.5 seconds. The task takes approximately 2 min. to complete.

##### *Digit Vigilance*

This task measures sustained and intensive attention; also known as vigilance. The participant is instructed to monitor a rapidly appearing series of digits presented one at a time in the center of the screen. At the start of the task a 'target' digit is selected randomly and presented on the right hand side of the screen where it remains throughout the 3 min. task. The digits are presented in an unpredictable order at the rate of 150 per min., and there are 15 targets every minute. The participant is instructed to press the RIGHT arrow keyboard key as quickly as possible every time a target digit appears in the series of digits, even if the digit is no longer displayed. As in simple reaction time, this is the only type of response made during the task, and the participant is again required to keep the forefinger lightly on the key throughout the test. The task records the number of correct detections (hits), the speed of these correct detections, and all responses made in error (false alarms). By having a high target rate, the task effectively monitors the quality of focus second by second over the 3 min. task duration. The advantage of having the target digit constantly displayed is that it removes any involvement of working memory from the task, allowing the quality of performance of the task to be a pure measure of attentional ability.

##### *Choice Reaction Time*

This task measures alertness, the ability to focus concentration as well as information processing. Besides being a measure of attention and focus, this task also measures the processing time required to identify the stimulus and select the appropriate response. The task is similar to Simple Reaction Time with the exception that each stimulus can be either the word YES or the word NO. The participant is asked to place the left forefinger on the LEFT arrow keyboard key and the right forefinger on the RIGHT arrow keyboard key. They are instructed that stimuli will be presented in the centre of the screen in an unpredictable order. The participant is instructed to press the RIGHT arrow keyboard key whenever the word YES is presented, or the LEFT arrow keyboard key whenever the word NO is presented. The participant is instructed to press the appropriate key as quickly and accurately as possible. It is emphasized that the fingers should remain on the buttons throughout the task. 50 stimuli are used with randomly varying intervals between 1 and 3.5 seconds. The task records speed and accuracy of all responses and takes approximately 2 min. to complete.

#### Working Memory & Executive Control

##### *Spatial Working Memory*

This task measures the ability to keep spatial information in working memory and to retrieve it. The participant is presented with a 3x3 array of light bulbs for 10 seconds. Four are lit, and the participant must remember the spatial location/pattern of these bulbs. A series of 'probe' stimuli (3 x 3 arrays) in which only one bulb is lit, is shown on the screen one at a time, each lit bulb position of the original target pattern is probed four times (16) and each non-target position is probed four times (20) giving 36 probes in total. The order of presentation is randomized. During this part of the task the participant is instructed to place the left forefinger on the left arrow keyboard key, and the right forefinger on the right arrow keyboard key. The participant is instructed to press the RIGHT arrow keyboard key whenever an original lit bulb is presented, or the LEFT arrow keyboard key if the probe is not one of the original items. The probes remain on the screen until the key is pressed, and the participants are instructed to make their decisions as quickly and accurately as possible. The accuracy of responses is recorded, as is the speed of all appropriate responses. The task lasts approximately 1.5 min.

##### *Numeric Working Memory*

This task measures a participant's ability to hold numeric information in working memory and rapidly retrieve it. A 'target' series of 5 digits (0 to 9) is presented one at a time. Each digit is displayed for 1150 ms with an interval of 50 ms between each presentation. A series of 30 probe digits follow (digits from 0-9), and the participant is instructed to press the RIGHT arrow keyboard key if the digit was one of the targets originally presented, or the LEFT arrow keyboard key if it was not. Probe stimuli remain on screen until a response is made. Half of the probe stimuli require a RIGHT arrow key response and half a LEFT arrow key response. During this part of the task the participant is instructed to place the left forefinger on the LEFT arrow keyboard key, and the right forefinger on the RIGHT arrow keyboard key, and to make the responses as quickly and accurately as possible. The task last for approximately 1.5 min.

#### Episodic/Declarative Memory

##### *Verbal Recall & Recognition*

This task measures episodic memory for verbal information. Fifteen words are initially presented on screen, one at a time; each word is displayed for 1500 ms with an interval of 500 ms between each presentation (one word every 2 seconds). Immediate word recall follows this task directly after the final word has been presented; recall is achieved by participants typing as many words as they can recall (in any order) within 60 seconds. Participants then complete the CogTrack™ tests of attention and working memory, followed by delayed word recall and delayed word recognition. In delayed word recall, the participant again types as many words as they can remember seeing earlier (in any order) within 60 seconds. Word Recognition then follows, in which the original words together with a number of equivalent distractor words are presented one at a time, in a random order. During this part of the task the participant is instructed to place the left forefinger on the LEFT arrow keyboard key, and the right forefinger on the RIGHT arrow keyboard key. The participant is instructed to press the RIGHT arrow keyboard key whenever an original word is presented, or the LEFT arrow keyboard key if it is not one of the original words. It is emphasized that the responses must be made as quickly and accurately as possible. Each word remains on the screen until a response is made and the speed and accuracy of every response is recorded. A different but equivalent list of words is presented on each testing occasion. These tasks take approximately 4 min. in total.

##### *Picture Recognition*

This task measures the ability to store and retrieve visual information, being a measure of cued episodic secondary memory retrieval. A series of 20 pictures of everyday scenes and objects is presented on the screen at the rate of 1 every 3 seconds. The participant is instructed to pay close attention to the detail of each picture, as they will later be shown these pictures together with very similar ones (named lures). There are no responses for this part of the task. Then, after other intervening tests have been performed, usually around 15 min. later, the 20 original pictures are presented mixed with the 20 lure pictures. During this part of the task the participant is instructed to place the left forefinger on the left arrow keyboard key, and the right forefinger on the right arrow keyboard key. Each picture has a closely similar paired picture, and the participant is instructed to press the RIGHT arrow keyboard key whenever an original picture is presented, or the LEFT arrow keyboard key if it is a different one. Again the stimuli remain on the screen until the key is pressed, and the participant is instructed to make the decisions as quickly and accurately as possible. The accuracy and speed of every response is recorded. The two parts of the task together take approximately 3 min. and 30 seconds. Different picture pairs are used for each test run. The accuracy and speed of response to the lure stimuli in this task have been shown to be selectively sensitive to activity in the dentate gyrus of the hippocampus, the area responsible for neurogenesis.

#### Procedure

##### Visit 1 (V1)

During V1 (approximately 45 mins), which followed telephone screening, inclusion/exclusion criteria were checked and documented by a study physician, including medical history and vital signs measurements (BMI, WHR, BFP, MP, pulse, BP) demographic data of eligible individuals were assessed and an appointment for V2 (maximum 7 days after V1) was arranged. Participants were instructed not to consume any alcohol until the next visit. Additionally, the last meal intake should be 1h before V2 and participants were instructed to drink only water for one hour before V2. Females of childbearing potential who do not use contraceptives were instructed to refrain from heterosexual intercourse for the entire study duration.

##### Visit 2 (V2)

During V2 (approximately 60 mins), participants were asked about their medical history, intake of concomitant medication, their sleep duration and quality, their lifestyle and level of activity as well as their alcohol consumption. Participants filled in the TICS and underwent two training sessions for familiarization with the cognitive tests. A saliva collection kit containing three Salivettes® (Sarstedt, Nuembrecht, Germany) for assessing the CAR on the third study day was handed out and participants were instructed not to consume any alcohol. Additionally, the last meal intake should be 1h before V3 and participants were instructed to drink only water for one hour before V3, which was scheduled on the next day.

##### Visit 3 (V3)

On the morning of V3 participants were asked to collect saliva samples at home immediately upon awakening, and then subsequently 30 minutes and 45 minutes after awakening. Participants were instructed not to eat or brush their teeth during the collection, but were allowed to drink water until 5 minutes before each saliva sample.

V3 took place between 12 noon and 3pm (starting time) approximately 24 hours after V2. Each participant's appointment lasted approximately 240 min. Adverse Events were recorded as they occurred during V3 after study product intake.

65 minutes prior to product administration (-65 minutes) participants were given a detailed introduction to the study procedure of V3, and asked about their medical history, intake of concomitant medication, their sleep duration and quality, and their alcohol consumption. Vital signs (BP, pulse rate) were assessed. Participants were instructed to drink only water and not to eat during V3 besides the standard meal provided.

At -35 minutes saliva sample #1 was collected, and participants BP and HR were assessed.. Baseline cognitive tests were then performed on CogTrack™ at -25 minutes and following this, saliva sample #2 was collected, and participants BP and HR were assessed at -5 minutes.

Product administration occurred according to the randomisation plan, and at +50 minutes saliva sample #3 was collected, and participants BP and HR were assessed. At +60 minutes cognitive tests were performed (CogTrack™, 1h post study product administration) and following this (+80 minutes) saliva sample #4 was collected, and participants BP and HR were assessed.

Subjects received a standardised meal at +90 minutes. At +140 minutes saliva sample #5 was collected, and participants BP and HR were assessed, and then at +150 minutes cognitive tests were performed (CogTrack™, 2.5hrs post study product administration). Following this (+170 minutes), saliva sample #6 was collected, and participants BP and HR were assessed.
